# The dilution effects of healthy lifestyles on the risk of depression attributed to life-course disadvantages among Chinese middle-aged and older adults

**DOI:** 10.1101/2022.09.14.22279926

**Authors:** Ziyang Ren, Yushan Du, Xinyao Lian, Xiaoying Zheng, Jufen Liu

**Author notes:** Correspondence: Jufen Liu, PhD, Institute of Reproductive and Child Health / Key Laboratory of Reproductive Health, National Health Commission of the People’s Republic of China; Department of Epidemiology and Biostatistics, School of Public Health, Peking University, Beijing 100191, China. Xiaoying Zheng, School of Population Medicine and Public Health, Chinese Academy of Medical Sciences/Peking Union Medical College, Beijing, China.

## Abstract

**Background:** Life-course disadvantages and unhealthy lifestyles are well-known to independently induce depression, but their joint effects on depression and whether adopting healthy lifestyles can dilute the depressive risks attributed to life-course disadvantages remain unknown in China.

**Methods:** Data were from the China Health and Retirement Longitudinal Study. Life-course (childhood and adulthood) disadvantages were defined using 60 questions and were classified as mild, moderate, and severe by principal component analysis. Healthy lifestyles, including regular exercise, reasonable sleep, never smoking, and no heavy alcohol consumption, were classified as 0/1, 2, 3, and 4. Depression was assessed using CESD-10 with cutoff ≥10. Logistic regression was used to investigate the association of life-course disadvantages and lifestyles with depression and the dilution effects of healthy lifestyles on the depressive risks attributed to life-course disadvantages.

**Results:** Among 5724 participants included, sleep and smoking mediated 5.1% and 1.1% the effect of severe life-course disadvantages on depression while exercise mediated - 0.8%. Multiple (vs. 0/1) healthy lifestyles were associated with decreased depressive risks more significantly as the life-course disadvantages increased, with odds ratios (ORs) and 95% confidence intervals (CIs) of 0.44 (0.25-0.80) and 0.33 (0.21-0.53) for 4 healthy lifestyles in participants with mild and severe life-course disadvantages, respectively. Compared to participants with mild life-course disadvantages and 4 healthy lifestyles, those who with severe life-course disadvantages and less than one lifestyles were more than 10 times as significantly associated with depression. Finally, participants with mild/moderate life-course disadvantages but ≤2 healthy lifestyles (vs. severe but 4 lifestyles) were not associated with decreased depressive risks.

**Conclusion:** Adopting multiple healthy lifestyles can well-dilute the depressive risks attributed to life-course disadvantages in middle-aged and older Chinese, which is of great importance for reducing the depressive burden and the construction of healthy aging in China.

## Introduction

Depression, a common disorder that severely impairs psychosocial functioning and lowers the quality of life, was reported to affect 39 % of the elderly (65+) and 45% of the most elderly (80+) in China^1,2^. The scientific community has recently highlighted the importance of preventing depression, especially in the senior Chinese^2-4^. Healthy lifestyles, given their high cost-effectiveness, have been recognized as one of the most practical primary preventatives against depression^5,6^.

Life-course disadvantages refer to a wide spectrum of stressful socioeconomic, interpersonal, mental, and health-related conditions across the life-course, including those occurring in both childhood and adulthood^7-9^. Individuals with life-course disadvantages are more likely to experience depression and develop unhealthy life habits, as has been well demonstrated^10-13^. However, given the stigma of mental illness and cultural differences^14^, related evidence in the Chinese population is much less adequate than that in western populations.

Limited evidence, to the best of our knowledge, has comprehensively demonstrated the association of life-course disadvantages and various lifestyles with depression in China. According to earlier research, sleep quality mediated the association between disadvantages and depression^15,16^. Boisgontier, et al. also indicated that physical activity reduced the effect of adverse childhood experiences on depression^17^. However, their studies were constrained due to the insufficient definition of life-course disadvantages and the limited variety of healthy lifestyles. Additionally, given the stark differences between Chinese and Western lifestyle habits and the heavy burden of depression in the elderly Chinese^3^, it is worth exploring whether adopting healthy lifestyles can assist Chinese seniors who are at high risk for depression, but few studies have been conducted to date.

To fill in these knowledge gaps, we utilized the China Health and Retirement Longitudinal Study (CHARLS) to investigate the association of life-course disadvantages with depression and the role of unhealthy lifestyles and whether adopting healthy lifestyles can dilute the depressive risks attributed to life-course disadvantages in middle-aged and older Chinese.

## Method

### Study Population

Data were from the 2014 and 2018 China Health and Retirement Longitudinal Study (CHARLS), a nationally representative survey that included Chinese aged 45 years and older from 450 villages/urban communities in 28 provinces across China using a stratified multistage probability sampling strategy. The detailed procedures of the CHARLS have been reported elsewhere^18^. The baseline survey was conducted in 2011, with follow-ups in 2013, 2015, and 2018, respectively. In 2014, the CHARLS further carried out a life history survey to retrospectively record the respondents’ life experiences from birth. The CHARLS was approved by the Ethical Review Committee of the Peking University and implemented by the China Centre for Economic Research of Peking University. All participants provided written informed consent.

Among 17324 participants recruited in both 2014 and 2018, those who aged < 45 or with missing data on sex (N=445); who had missing data on lifestyles and No. of diseases (N=102); who had missing data on any childhood disadvantages (N=8003); who had missing data on residence, household income, and other adulthood disadvantages (N=2423); and who had missing data on depression (N=627) were excluded, leaving 5724 participants for our main analysis **(Figure S1)**.

### Definition of life-course disadvantages

The detailed definitions of life-course disadvantages are shown in **Table S1**. All questions related to childhood disadvantages and adulthood adversities were asked through questionaries in 2014, while information on adulthood socioeconomic status (SES), like household income and residence was collected in 2018. According to the previous research^19^, 48 childhood disadvantages, including 8 items in childhood SES, 6 in childhood health, 2 in childhood wars, 17 in childhood traumas, and 15 in childhood relationships, as well as 12 adulthood disadvantages, including 4 items in adulthood SES and 8 in adulthood adversities, were defined in the CHARLS. Life-course disadvantages were calculated as the sum of childhood and adulthood disadvantages, for a total of 60 items.

We further employed the principal component analysis (PCA) for all the items of life-course, childhood, or adulthood disadvantages, respectively, and selected components with eigenvalues of >2 to cluster participants into 3 equal parts: mild, moderate, and severe. The eigenvalues and percentages of explained variances of the first 10 components are shown in **Figure S2 and Figure S3**. We used the first 3, 2, and 1 principal components of life-course, childhood, and adulthood disadvantages, accounting for 15.74%, 12.47%, and 19.92% of the variances, respectively.

### Definition of healthy lifestyles

We only included 4 healthy lifestyles, including regular exercise, reasonable sleep, never smoking, and no heavy alcohol consumption in this study given the CHARLS did not gather information on dietary intake.

The duration of exercise was calculated by multiplying the weekly frequency and the duration of each session according to prior research^20^. Regular exercise was defined as at least 150 minutes of moderate-intensity or 75 minutes of vigorous-intensity physical activity per week as recommended by the 2020 guidelines of the World Health Organization (WHO)^21^. 7-8 hours of sleep duration each night was considered to be reasonable as recommended by China government^22^. Never smoking was defined if individuals had never smoked in their life-course. Heavy alcohol consumption was defined as > 14 drinks/week for males and > 7 drinks/week for females. A drink in China was 13.6 g of ethanol, which was equivalent to 375 ml of beer or 118 ml of wine^23^. Healthy lifestyles were then calculated as the sum of regular exercise, reasonable sleep, never smoking, and no heavy alcohol consumption, ranging from 0-4. Given the limited number of participants with no healthy lifestyle, we further classified healthy lifestyles as 0/1, 2, 3, and 4.

### Definition of Depression

The 10-item Center for Epidemiological Studies Depression Scale (CESD-10)^24^ was used to define depression. Participants were asked “how you have felt and behaved during the last week”, with options including 1) rarely or none of the time (one day); 2) some or a little of the time (1-2 days); 3) sometimes or a significant amount of the time (3-4 days); and 4) most or all of the time (5-7 days). These answers were sequentially assigned as 0-3, for a total score of 0 to 30. Depression was then defined as a score ≥10^25,26^.

### Covariates

Socioeconomic and health-related data like age, sex, and No. of diseases were collected through face-to-face interviews. The No. of diseases in this study was defined as the sum of self-reported main chronic diseases, such as diabetes, hypertension, dyslipidemia, stroke, and heart diseases, and was classified as 0, 1, and 2 or more.

### Statistical Analysis

The baseline characteristics of included participants were described as medians with interquartile ranges (IQRs) for continuous variables, and frequency and percent (%) for categorical variables. To compare characteristics by life-course disadvantages, differences in continuous variables and categorical variables were assessed by Wilcoxon rank sum tests and Chi-square tests.

We utilized logistic regression to examine the associations of life-course, childhood, and adulthood deficits with depression based on the PCA results given above. Models for life-course disadvantages were first adjusted for age, sex, and No. of diseases and then further adjusted for lifestyles, whereas those for childhood or adulthood disadvantages were additionally adjusted for adulthood or childhood disadvantages first. The ‘mediation’ package, Poisson regression, and Logistic regression were used to determine the mediating role of lifestyles. For instance, the following equations were used for childhood disadvantages.

*Model* 1: *No. of healthy lifestyles* = *childhood disadvantages* +*adulthood disadvantages* + *age* + *sex* + *No. of diseases* (*Poisson regression*)

*Model* 2: *Depression* = *childhood disadvantages* + *No. of healthy lifestyles* + *adulthood disadvantages* +*age* + *sex* + *No. of diseases* (*Logistice regression*)

After further adjusting for other lifestyles, we investigated the mediating role of specific lifestyles. Instead of utilizing Poisson regression, Model 1 for specific lifestyles in mediation analysis was conducted using Logistic regression.

Additionally, the associations between No. of healthy lifestyles and depression in the total population, males, and females were explored after adjusting for age, sex (except sex-stratified analysis), No. of diseases, and life-course disadvantages. Models for specific healthy lifestyles were further adjusted for other lifestyles. We also investigated the association between No. of healthy lifestyles and depression in participants with mild, moderate, and severe life-course, childhood, and adulthood disadvantages, respectively, with the reference given as 0/1. Lifestyle-specific and sex-stratified analyses were also conducted.

The joint associations of life-course, childhood, or adulthood disadvantages and unhealthy lifestyles with depression were investigated in the total population, males, and females. The reference was assigned as mild and 4 healthy lifestyles. Models for life-course disadvantages were adjusted for age, sex (except sex-stratified analysis), and No. of diseases, whereas models for childhood/adulthood disadvantages were further adjusted for adulthood/childhood disadvantages.

After adjusting for age, sex, and No. of diseases for life-course disadvantages; adulthood disadvantages, age, sex, and No. of diseases for childhood disadvantages; and childhood disadvantages, age, sex, and No. of diseases for adulthood disadvantages, we explored the dilution effects of No. of healthy lifestyles on the association of life-course, childhood, or adulthood disadvantages with depression. The reference was assigned as severe and 4 healthy lifestyles. Lifestyle-specific analyses were additionally adjusted for other lifestyles. Sex-stratified analyses were also conducted.

Reporting of this study was done under Strenghtening the Reporting of Observational studies in Epidemiology (STROBE) guidelines^27^. Analyses were performed using SAS statistical software version 9.4 (SAS Institute) and R statistical software version 4.1.2 (R Project for Statistical Computing). All analyses were two-sided, and a *P* value of < 0.05 and a 95% confidence interval (CI) of odds ratio (OR) that did not cross 1.00 was considered statistically significant.

## Results

The baseline characteristics of included participants are shown in **Table S2**. The age of participants with mild, moderate, and severe life-course disadvantages is 58.0 (53.0-66.0), 61.0 (53.0-67.0), and 61.0 (53.0-68.0). We found significant differences in age, sex, regular exercise, reasonable sleep, never smoking, no heavy alcohol consumption, No. of diseases, depression, childhood disadvantages, and adulthood disadvantages among participants with different life-course disadvantages (all *P* values <0.05).

**Table 1** demonstrates that moderate and severe (vs. mild) life-course, childhood, and adulthood disadvantages are significantly associated with depression, with fully adjusted ORs (95% CIs) of 1.59 (1.34-1.88) and 3.33 (2.84-3.91) for moderate and severe life-course disadvantages, 1.46 (1.23-1.73) and 2.48 (2.10-2.93) for moderate and severe childhood disadvantages, and 1.50 (1.25-1.79) and 2.67 (2.26-3.15) for moderate and severe adulthood disadvantages. It was also observed that lifestyles mediated about 3.1% and 3.8% the effects of severe life-course and adulthood disadvantages on depression, respectively. More precisely, we discovered that sleep mediated 5.1%, 5.0%, and 2.5% the effect of severe life-course, childhood, and adulthood disadvantages on depression and that smoking mediated 1.1% and 1.9% the effect of severe life-course and adulthood disadvantages on depression. Interestingly, we found that exercise played a negative mediating role in the associations of life-course, childhood, and adulthood disadvantages with depression **(Table S3)**.

**Table 1.** Associations of life-course disadvantages with depression, and the mediation proportion attributed-lifestyles

|  | Mild | Moderate | Severe |
| --- | --- | --- | --- |
|  | OR (95% CI) |  |  |
| *Life-course disadvantages* |  |  |  |
| Unadjusted for lifestyles | Reference | **1.59 (1.35-1.88)** | **3.43 (2.92-4.02)** |
| Adjusted for lifestyles | Reference | **1.59 (1.34-1.88)** | **3.33 (2.84-3.91)** |
| Mediation proportion (%) (95% CI) | - | 3.0 (-5.4 to 10.2) | **3.1 (0.5 to 6.0)** |
| *P* value | - | 0.420 | 0.018 |
| *Childhood disadvantages* |  |  |  |
| Unadjusted for lifestyles | Reference | **1.47 (1.24-1.73)** | **2.49 (2.11-2.94)** |
| Adjusted for lifestyles | Reference | **1.46 (1.23-1.73)** | **2.48 (2.10-2.93)** |
| Mediation proportion (%) (95% CI) | - | 0.1 (-10.6 to 9.9) | 1.0 (-2.7 to 4.9) |
| *P* value | - | 0.990 | 0.580 |
| *Adulthood disadvantages* |  |  |  |
| Unadjusted for lifestyles | Reference | **1.51 (1.26-1.80)** | **2.75 (2.33-3.24)** |
| Adjusted for lifestyles | Reference | **1.50 (1.25-1.79)** | **2.67 (2.26-3.15)** |
| Mediation proportion (%) (95% CI) | - | 2.5 (-6.3 to 11.8) | **3.8 (0.5 to 7.4)** |
| *P* value | - | 0.520 | 0.022 |

Notes: OR, odds ratio. CI, confidence interval. Models for life-course disadvantages were first adjusted for age, sex, and No. of diseases and then further adjusted for lifestyles. Models for childhood disadvantages were first adjusted for adulthood disadvantages, age, sex, and No. of diseases and then further adjusted for lifestyles. Models for adulthood disadvantages were first adjusted for childhood disadvantages, age, sex, and No. of diseases and then further adjusted for lifestyles. Bolded indicates statistical significance.

Furthermore, adopting healthy lifestyles was associated with decreased depressive risks in both males and females (OR=0.26, 95% CI: 0.12-0.57, for 4 healthy lifestyles in males; OR=0.31, 95% CI: 0.19-0.51, for 4 healthy lifestyles in females when compared to 0/1 healthy lifestyle) though not all ORs (95% CIs) of specific lifestyles were statistically significant (**Table S4 and Table S5)**. According to **Figure 1**, we found that the negative associations between No. of healthy lifestyles and depression were more significant as life-course, childhood, and adulthood disadvantages increased, whereas the interactive effects between either disadvantage and healthy lifestyles were not observed (all *P* values for the interaction >0.05). Findings of sex-stratified analyses were in line with our main results **(Table S6)**. In participants with severe life-course disadvantages, the lifestyle-specific analysis revealed that regular exercise, reasonable sleep, and never smoking, but not no heavy alcohol consumption, were negatively associated with depression, with ORs (95% CIs) of 0.77 (0.63-0.93) for regular exercise, 0.61 (0.49-0.76) for reasonable sleep, and 0.70 (0.53-0.93) for never smoking. These findings were also observed in sex-stratified analyses **(Table 2 and Table S7)**.

**Table 2.** Association of specific healthy lifestyles with depression by life-course disadvantages

| Specific healthy lifestyles | Mild | Moderate | Severe | P for interaction |
| --- | --- | --- | --- | --- |
|  | OR (95% CI) |  |  |  |
| *Life-course disadvantages* |  |  |  |  |
| Regular exercise | 0.97 (0.75-1.26) | 0.92 (0.73-1.16) | **0.77 (0.63-0.93)** | 0.489 |
| Reasonable sleep | **0.54 (0.41-0.71)** | **0.61 (0.48-0.78)** | **0.61 (0.49-0.76)** | 0.618 |
| Never smoking | **0.59 (0.39-0.88)** | 0.74 (0.52-1.05) | **0.70 (0.53-0.93)** | 0.644 |
| No heavy alcohol consumption | 1.03 (0.70-1.51) | 0.85 (0.64-1.13) | 0.85 (0.64-1.13) | 0.885 |
| *Childhood disadvantages* |  |  |  |  |
| Regular exercise | 0.99 (0.76-1.28) | 0.86 (0.69-1.07) | **0.77 (0.62-0.94)** | 0.453 |
| Reasonable sleep | **0.54 (0.41-0.71)** | **0.63 (0.50-0.80)** | **0.60 (0.48-0.75)** | 0.547 |
| Never smoking | 0.66 (0.44-1.00) | 0.74 (0.53-1.03) | **0.74 (0.55-0.99)** | 0.863 |
| No heavy alcohol consumption | 1.00 (0.69-1.45) | 0.86 (0.64-1.14) | 0.93 (0.73-1.18) | 0.717 |
| *Adulthood disadvantages* |  |  |  |  |
| Regular exercise | 0.91 (0.70-1.20) | 0.80 (0.63-1.01) | 0.88 (0.72-1.06) | 0.690 |
| Reasonable sleep | **0.47 (0.35-0.64)** | **0.69 (0.54-0.88)** | **0.60 (0.49-0.73)** | 0.204 |
| Never smoking | **0.63 (0.42-0.96)** | 0.89 (0.62-1.30) | **0.68 (0.52-0.90)** | 0.058 |
| No heavy alcohol consumption | 1.25 (0.87-1.80) | **0.73 (0.53-0.99)** | 0.92 (0.73-1.16) | 0.689 |

Notes: OR, odds ratio. CI, confidence interval. Models for life-course disadvantages were adjusted for age, sex, No. of diseases, and other lifestyles. Models for childhood disadvantages were adjusted for adulthood disadvantages, age, sex, No. of diseases, and other lifestyles. Models for adulthood disadvantages were adjusted for childhood disadvantages, age, sex, No. of diseases, and other lifestyles. Bolded indicates statistical significance.

**Figure 1.**
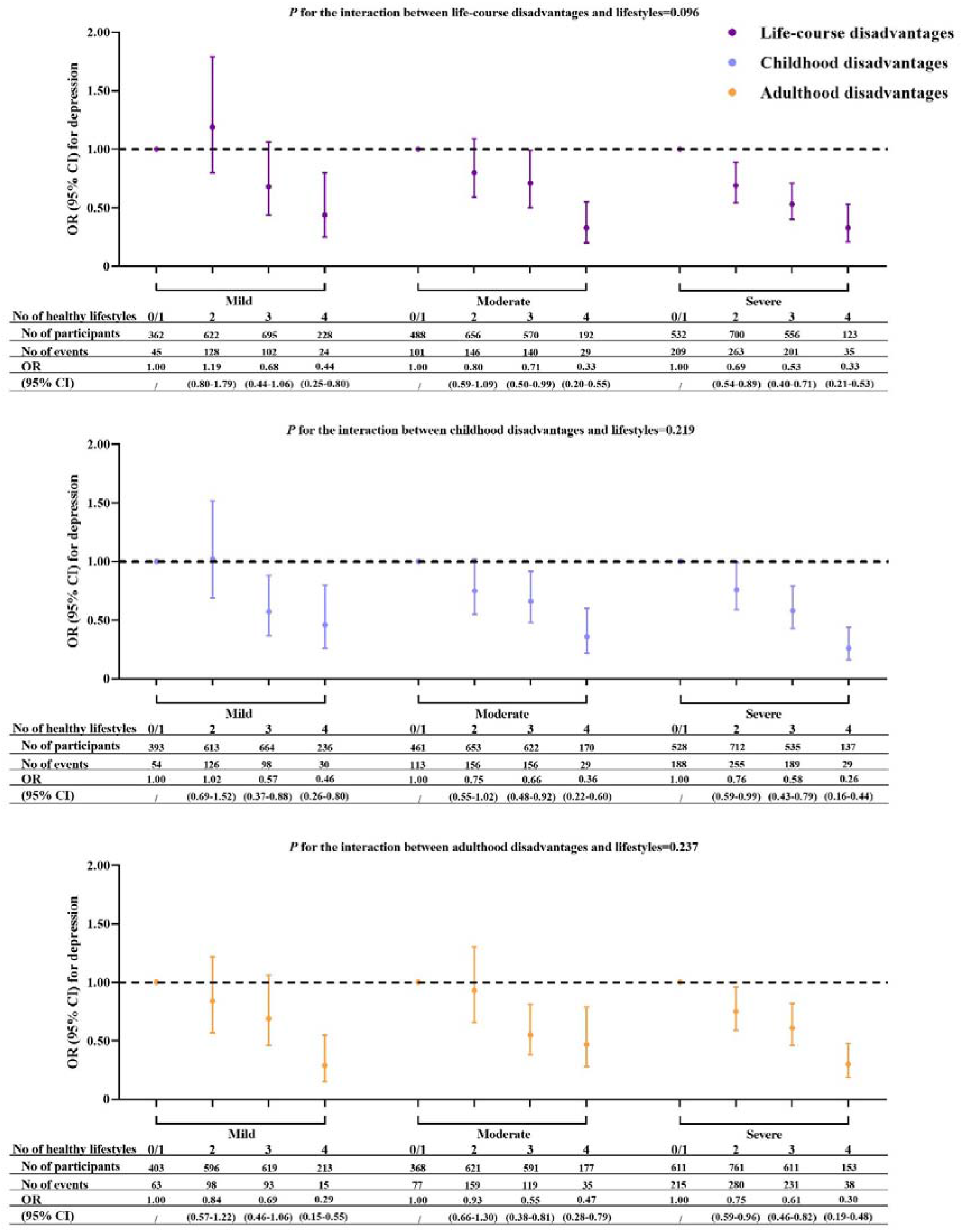
Association of healthy lifestyles with depression by life-course disadvantages Notes: OR, odds ratio. CI, confidence interval. Models for life-course disadvantages were adjusted for age, sex, and No. of diseases. Models for childhood disadvantages were adjusted for adulthood disadvantages, age, sex, and No. of diseases. Models for adulthood disadvantages were adjusted for childhood disadvantages, age, sex, and No. of diseases.

Figure 2. shows the joint associations of life-course, childhood, or adulthood disadvantages and unhealthy lifestyles with depression. Participants were more likely to experience depression if they had more severe disadvantages and adopted fewer healthy lifestyles. Similar results of sex-stratified analyses are shown in **Table S8**. Subsequently, we found that participants with moderate life-course disadvantages and 3 or less healthy lifestyles, as well as those with mild life-course disadvantages and 2 or less healthy lifestyles, were not at significantly higher depressive risks when compared to those with severe life-course disadvantages and 4 healthy lifestyles **(Figure 3)**. Meaningfully, we found that participants with moderate childhood disadvantages and 0/1 healthy lifestyle (vs. severe childhood disadvantages and 4 healthy lifestyles) were more likely to experience depression. We also conducted sex-stratified analyses and found similar results **(Table S9)**. When investigating the dilution effects of specific healthy lifestyles, we did not find any substantial dilution effects of specific healthy lifestyles on the depressive risks caused by life-course disadvantages. However, reasonable sleep and never smoking may dilute the depressive risks caused by childhood and adulthood disadvantages. The dilution effects of reasonable sleep and never smoking on the depressive risks attributed to life-course disadvantages were also verified in both males and females **(Table S10 and Table S11)**.

**Figure 2.**
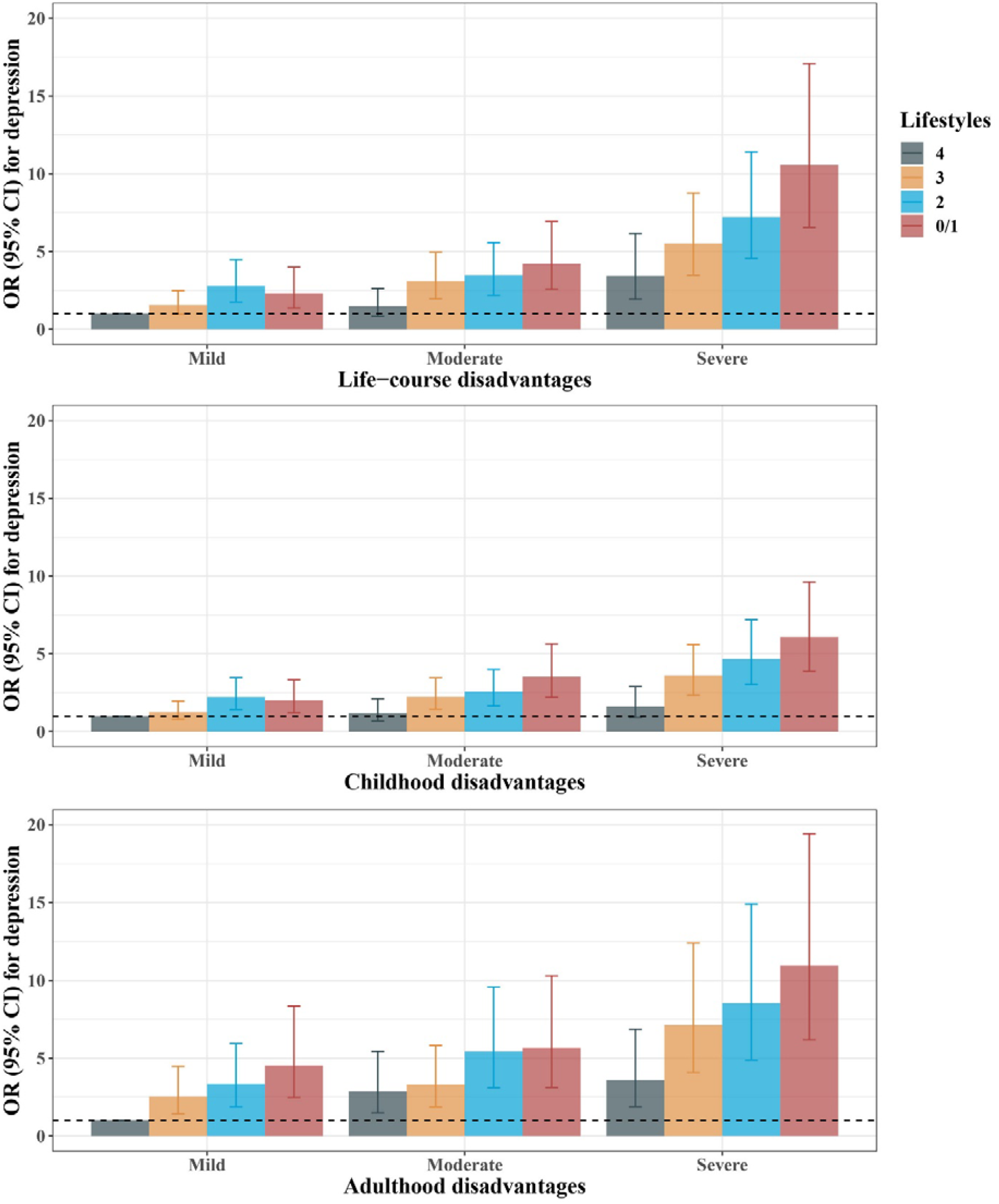
Joint association of life-course disadvantages and unhealthy lifestyles with depression Notes: OR, odds ratio. CI, confidence interval. Models for life-course disadvantages were adjusted for age, sex, and No. of diseases. Models for childhood disadvantages were adjusted for adulthood disadvantages, age, sex, and No. of diseases. Models for adulthood disadvantages were adjusted for childhood disadvantages, age, sex, and No. of diseases.

**Figure 3.**
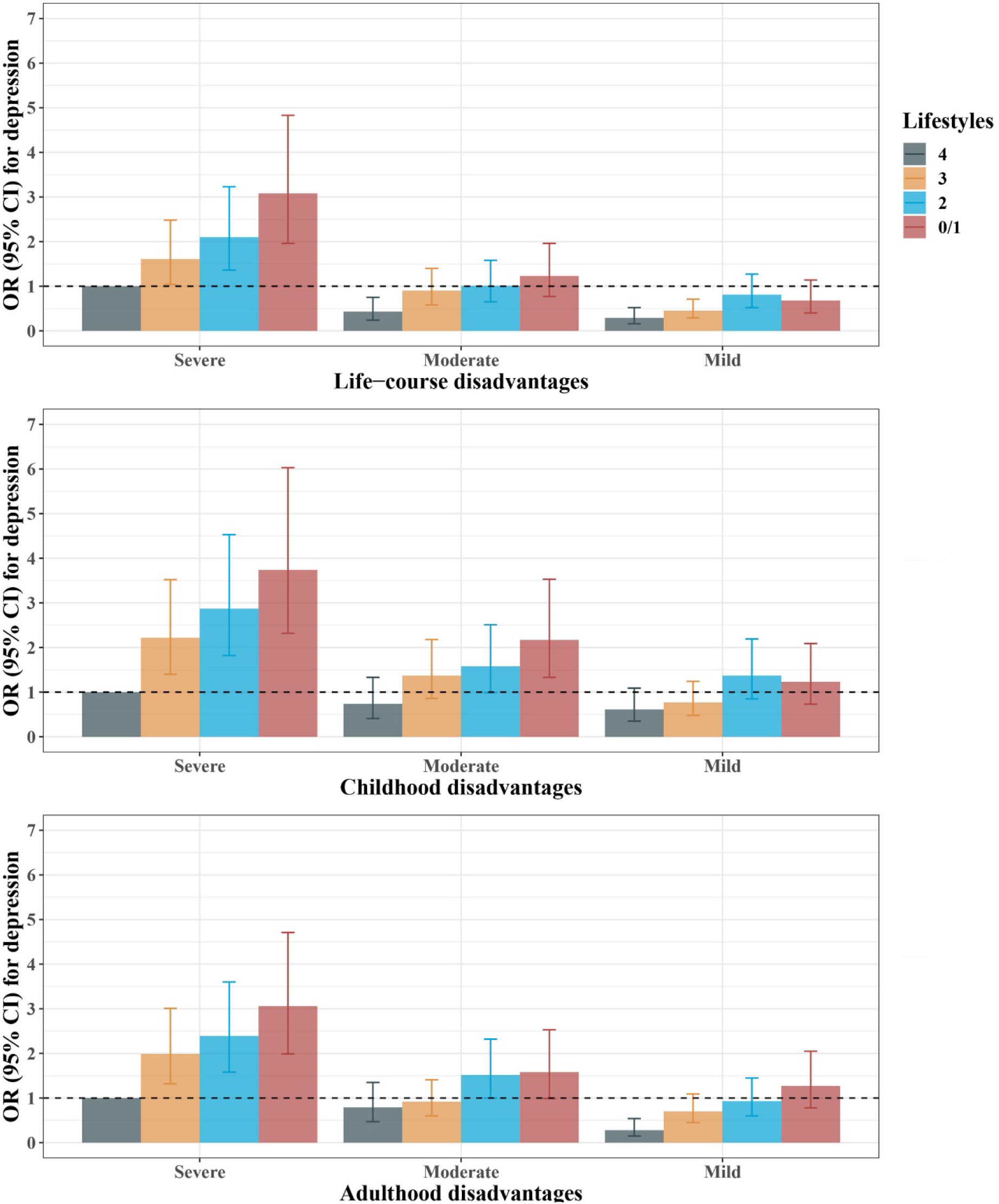
The dilution effect of No. of healthy lifestyles on the association between life-course disadvantages and depression Notes: OR, odds ratio. CI, confidence interval. Models for life-course disadvantages were adjusted for age, sex, and No. of diseases. Models for childhood disadvantages were adjusted for adulthood disadvantages, age, sex, and No. of diseases. Models for adulthood disadvantages were adjusted for childhood disadvantages, age, sex, and No. of diseases.

## Discussion

In this retrospective study, we found growing associations of multiple healthy lifestyles with lower depressive risks as life-course disadvantages increased and substantial joint effects of life-course disadvantages and unhealthy lifestyles on depression in middle-aged and older Chinese. More importantly, adopting multiple healthy lifestyles was found to dilute the risks of depression attributed to life-course disadvantages and even mask some of the depressive risks caused by childhood disadvantages. We also found that sleep and smoking positively mediated the effect of life-course disadvantages on depression, whereas exercise negatively mediated this effect.

According to the baseline characteristics across life-course disadvantages, we found that participants with more severe life-course disadvantages were more likely to engage in regular exercise but less likely to get reasonable sleep and never smoking, which can be used to interpret the negative mediating role of exercise and the positive mediating role of sleep and smoking. Numerous studies have shown that stressful life events are associated with substance abuse and poor sleep quality^28,29^. However, related studies in China have focused more on the effects of adverse childhood experiences^25,30^. Limited evidence has indicated that life-course disadvantages are associated with unreasonable sleep, ever smoking, and heavy alcohol consumption in middle-aged and older Chinese, as was verified in this study. In contrast to previous findings of non-significant or negative associations between trauma exposure and exercise^31,32^, we discovered that individuals with life-course disadvantages were more likely to adopt regular exercise. According to a prior meta-analysis, physical activity was positively impacted by stress given that some individuals may utilize exercise to cope with stress^33^, which may help explain the negative mediating role of exercise between life-course disadvantages and depression.

This study expanded on findings of earlier research that life-course disadvantages and unhealthy lifestyles were independently associated with an increased risk of depression^10,34,35^ and verified the joint effects of life-course disadvantages and multiple unhealthy lifestyles on depression. In addition, although prior evidence has demonstrated that adopting certain healthy lifestyles, such as regular exercise, can reduce the risks of depression associated with stressful events^17^, there have been few studies to investigate whether adopting multiple healthy lifestyles can dilute or even mask the depressive risks attributed to life-course disadvantages, as was confirmed in this study. According to earlier research, individuals with life-course stressful conditions and unhealthy lifestyles were more likely to develop high levels of chronic inflammation^36,37^. Increased inflammatory levels, on the other hand, are widely considered one of the main contributors to depression^38^. Hence, the tremendous joint effects of life-course disadvantages and unhealthy lifestyles and the dilution effects of healthy lifestyles on depression can be comprehended. Furthermore, either shorter or longer sleep is significantly associated with cognitive decline and metabolic abnormalities that are prevalent in the elderly and may further contribute to depression ^39-41^, which may help explain the most effective effects of reasonable sleep in our results. To the best of our knowledge, this is the first large-scale population-based study to investigate the association of life-course disadvantages and multiple lifestyles with depression in middle-aged and older Chinese. Our findings that different lifestyles may differently mediate the direction of life-course disadvantages’ effects on depression highlight the necessity of considering the impacts of each healthy lifestyle independently first when investigating the cumulative effects of healthy lifestyles, especially in senior Chinese. Additionally, we demonstrated that adopting healthy lifestyles can dilute or even mask the depressive risks attributed to life-course disadvantages, which provides a highly effective health intervention for individuals who are under stressful conditions and have a high risk of depression, assisting in alleviating the significant burden of depression among the elderly Chinese and thus promoting healthy aging. The rigorous implementation process and data collection methods of the CHARLS ensure the reliability of our findings.

### Limitations

Our study has several limitations. Given that dietary data were not gathered by the CHARLS, diet was not included in the lifestyles used in this study, but it will be explored in our future research. On the other hand, the participants included came from various regions of China, and their dietary habits and nutritional structure varied in a way that essentially did not affect the association between the four lifestyles used in this study and depression. In addition, information on life-course disadvantages was primarily self-reported, which may lead to potential recall bias. However, it’s a strategy that was widely used in most of the prior research. Finally, the cross-sectional design of this study cannot capture causal associations well, so our conclusions need to be further validated in subsequent large-scale cohort or intervention studies.

## Conclusion

In conclusion, we found that lifestyles mediated the effects of life-course disadvantages on depression; life-course disadvantages and unhealthy lifestyles had enormous joint effects on depression; and adopting multiple healthy lifestyles can dilute the depressive risks attributed to life-course disadvantages, in middle-aged and older Chinese. Our findings suggest that it is crucial to distinguish different lifestyle types when investigating their cumulative effects and provide representative population evidence to promote the widespread adoption of healthy lifestyles, especially 7-8 h of sleep per night, among middle-aged and elderly Chinese who are at high risk of depression, and the construction of healthy aging.

## Data Availability

All data produced are available online at http://charls.pku.edu.cn/

## Funding

Supported by the Fundamental Research Funds for the Central Universities (BMU2021YJ034) and the Major Project of the National Social Science Fund of China [grant numbers 21ZDA107; 21&ZD187].

## Conflicts of interest

The authors declare that they have no conflicts of interest.

## Authors’ contributions

JL designed the study. ZR managed and analyzed the data. ZR prepared the first draft. ZR, YD, and XL reviewed and edited the manuscript, with comments from JL and XZ. All authors were involved in revising the paper, JL had full access to the data and gave final approval of the submitted versions.

## Notes

### Competing Interest Statement

The authors have declared no competing interest.

