## Supplement for "The dilution effects of healthy lifestyles on the risk of depression attributed to life-course disadvantages among Chinese middle-aged and older adults"

**Table and Figure Legends**

### Table S1. Definition of life-course disadvantages in the CHARLS

| **Variables names** | | | **Questions** | **Responses/description** |
| --- | --- | --- | --- | --- |
| **Childhood disadvantages** | | |  |  |
|  | **Childhood SES** | |  |  |
|  |  | Parental education | What is the highest level of education your biological father/mother completed? | 0=both literate; 1=1 illiterate; 2=both illiterate |
|  |  | Parental occupation | What was your male guardian’s usual occupation when you were growing up before you were 17? | 0=non-farming; 1=farming |
|  |  | House type at birth | What is the architectural type of your first residence? | 1=concrete; 2=adobe; 3=wood or others |
|  |  | First hukou type | What was your first hukou type? | 0=agricultural; 1= non-agricultural |
|  |  | Family’s financial situation | When you were a child before age 17, compared to the average family in the same community/village at that time, how was your family’s financial situation? | 1=a lot/somewhat better; 2=same; 3=somewhat worse/a lot worse |
|  |  | Food deficiency | When you were a child before age 17 was there ever a time when your family did not have enough food to eat? | 0=no; 1=yes |
|  |  | Family starved to death | During those days, had any of your family (including your grandparents, parents, siblings, children and so on) starved to death? | 0=no; 1=yes |
|  |  | Parents political status | Is (Was) any of your parents a Communist Party member? | 0= yes; 1= no |
|  | **Childhood health** | |  |  |
|  |  | Self-reported health status | Before you were 15 years old (including 15 years old), would you say that compared to other children of the same age, you were healthier, same, or less healthy? | 1=healthier; 2=same; 3= less healthy |
|  |  | Ever confined to bed | Before you were 15 years old (including 15 years old), because of a health condition, were you ever confined to bed or home for a month or more? | 0=no; 1=yes |
|  |  | Ever hospitalized for 1 or more month | Before you were 15 years old (including 15 years old), because of a health condition, were you ever hospitalized for a month or more? | 0=no; 1=yes |
|  |  | Ever hospitalized more than 3 times | Were you ever hospitalized more than three times within a 12-month period before you were 15 years old (including 15 years old)? | 0=no; 1=yes |
|  |  | Received vaccinations | Before you were 15 years old (including 15 years old), have you received any vaccinations? | 0=yes; 1=no |
|  |  | Had a usual source of care | Before you were 15 years old (including 15 years old), have you always had a usual source of care, that is, a particular person or a place that you went to when you were sick or you needed advice about your health? | 0=yes; 1=no |
|  | **Childhood wars** | |  |  |
|  |  | Born in the Civil War era | Born in the Civil War era. (1946-1949) | 0=no; 1=yes |
|  |  | Born in the Second Sino-Japanese era | Born in the Second Sino-Japanese era. (1937-1945) | 0=no; 1=yes |
|  | **Childhood traumas** | |  |  |
|  |  | Parents died | Before age 16 did you one or both parents die? | 0=no; 1=yes |
|  |  | Parents divorced | Before age 16 did your biological parents divorce? | 0=no; 1=yes |
|  |  | Sibling died | Do you have any sibling who died before age 6? | 0=no; 1=yes |
|  |  | Parents had alcoholism problem | During the years you were growing up, did female/male dependents have alcoholism problem? | 0=no; 1=yes |
|  |  | Parents had smoking problem | During the years you were growing up, did female/male dependents have smoking problem? | 0=no; 1=yes |
|  |  | Parents had gambling problem | During the years you were growing up, did female/male dependents have gambling problem? | 0=no; 1=yes |
|  |  | Abused by neighbor kids | When you were a child, how often were you picked on or bullied by kids in your neighborhood? | 0=not very often/never; 1=often/sometimes |
|  |  | Abused by school kids | When you were a child, how often were you picked on or bullied by kids in your school? | 0=not very often/never; 1=often/sometimes |
|  |  | Hit by mother | When you were growing up, did your female guardian ever hit you? | 0=not very often/never; 1=often/sometimes |
|  |  | Hit by father | When you were growing up, did your male guardian ever hit you? | 0=not very often/never; 1=often/sometimes |
|  |  | Hit by siblings | When you were growing up, how often did your brother or sister ever hit you? | 0=not very often/never;1=often/sometimes |
|  |  | Parents ever quarreled | When you were growing up, how often did your parents ever quarrel? | 0=not very often/never; 1=often/sometimes |
|  |  | Parents hit each other | Have your parents hit each other? | 0=not very often/never; 1=often/sometimes |
|  |  | Parental sadness or depression | During the years you were growing up, had your female/male guardian showed continued signs of sadness or depression that lasted 2 weeks or more? | 0=no; 1=yes |
|  |  | Parents’ sick on bed | Did your female/male guardian have a long time be sick on bed when you were young? | 0=no; 1=yes |
|  |  | Parents’ deformity | Did your female/male guardian have a serious deformity when you were young? | 0=no; 1=yes |
|  |  | Parents’ abnormality of mind | Did your female/male guardian have abnormality of mind when you were young? | 0=no; 1=yes |
|  | **Childhood relationships** | |  |  |
|  |  | Self-rated neighborhood safety | Was it safe being out alone at night in the neighborhood where you lived as a child? | 0=very/somewhat; 1=not very/not at all |
|  |  | Self-rated neighborhood willingness to help | Were the neighbors of the place where you lived as a child willing to help each other out? | 0=very/somewhat; 1=not very/not at all |
|  |  | Self-rated neighborhood close-knit relationship | Were the neighbors of the place where you lived as a child very close-knit? | 0=very/somewhat; 2=not very/not at all |
|  |  | Self-rated neighborhood cleanness | Was the neighborhood of the place where you lived as a child very clean and attractive? | 0=very/somewhat; 3=not very/not at all |
|  |  | Felt lonely | When you were a child, how often did you feel lonely for not having friends? | 0=never/not very often; 1=sometimes/often |
|  |  | Had a group of friends | When you were a child, did you often have a group of friends that you felt comfortable spending time with? | 0=sometimes/often; 1=never/not very often |
|  |  | Had a good friend | When you were a child, did you have a good friend? | 0=yes; 1=no |
|  |  | Self-rated relationship with mother | How would you rate your relationship with your female guardian when you were growing up? | 1=excellent, 2= very good, 3=good, 4=fair, 5=poor |
|  |  | Mother’s love and affection | How much love and affection did your female guardian give you while you were growing up? | 0=often/sometimes; 1=rarely/never |
|  |  | Mother’s effort put into watching over you | How much effort did your female guardian put into watching over you? | 0=a lot/some; 1=a little/not at all |
|  |  | Mother’s preference for siblings | Did your female guardian treat your siblings better than you when you were growing up? | 0=a little / not at all; 1=very much/somewhat |
|  |  | Mother’s preference for boy | Did your female guardian prefer boys to girls? | 0=a little / not at all; 1=very much/somewhat |
|  |  | Self-rated relationship with father | How would you rate your relationship with your male guardian when you were growing up? | 1=excellent, 2= very good, 3=good, 4=fair, 5=poor |
|  |  | Father’s preference for siblings | Did your male guardian treat your siblings better than you when you were growing up? | 0=a little / not at all; 1=very much/somewhat |
|  |  | Father’s preference for boy | Did your male guardian prefer boys to girls? | 0=a little / not at all; 1=very much/somewhat |
| **Adulthood disadvantages** | | |  |  |
|  | **Adulthood SES** | |  |  |
|  |  | Individual’s education attainment | What is the highest level of education you attained now? | 1=college or more; 2=senior; 3=middle; 4=elementary; 5 illiteracy |
|  |  | Individual’s occupation | What is the main labor force status? | 1=others; 2=Non- agricultural employed;3=agricultural |
|  |  | Household income | Household income. | 1=third tertile; 2=second tertile; 3=first tertile |
|  |  | Residence | What is your residence: Rural or urban? | 0=urban; 1=rural |
|  | **Adulthood adversities** | |  |  |
|  |  | Child died | Death of child. | 0=no; 1=yes |
|  |  | Physically injury | Have you ever received a physical injury that has led to any permanent handicap, disability or limitations in what you can do in daily in adulthood? | 0=no; 1=yes |
|  |  | Experienced lifetime discrimination | After you were 16 years old, because of ill health, did you experience any of the following (Denied promotions, Assignment to a task with fewer responsibilities, working on tasks below your qualifications, Harassment by your boss or colleagues, Pay cuts, Dismissed)? | 0=no; 1=yes |
|  |  | Ever confined to bed | After you were 16 years old, because of a health condition, were you ever confined to bed or home for one month or more? | 0=no; 1=yes |
|  |  | Ever hospitalized for 1 or more month | After you were 16 years old, because of a health condition, were you ever hospitalized for a month or more? | 0=no; 1=yes |
|  |  | Ever hospitalized 3 or more times | Were you ever hospitalized more than three times within a 12- month period after you were 16 years old? | 0=no; 1=yes |
|  |  | Ever left job for health condition | After you were 16 years old, because of a health condition, did you leave your job for one month or more? | 0=no; 1=yes |
|  |  | Had a usual source of care | When you were between 16-55 years old, have you always had a usual source of care, that is, a particular person or a place that you went to when you were sick or you needed advice about your health? | 0=yes; 1=no |

Notes: CHARLS, China Health and Retirement Longitudinal Study. SES, socioeconomic status.

### Table S2. Baseline characteristics of included participants by life-course disadvantages

| **Baseline Characteristics** | | **Mild (N=1907)** | **Moderate (N=1906)** | **Severe (N=1911)** | ***P* value** |
| --- | --- | --- | --- | --- | --- |
| Age, year | | 58.0 (53.0-66.0) | 61.0 (53.0-67.0) | 61.0 (53.0-68.0) | <0.001 |
| Sex | |  |  |  | <0.001 |
|  | Male | 917 (48.1) | 1120 (58.8) | 1155 (60.4) |  |
|  | Female | 990 (51.9) | 786 (41.2) | 756 (39.6) |  |
| Regular exercise | |  |  |  | <0.001 |
|  | Yes | 1041 (54.6) | 1125 (59.0) | 1160 (60.7) |  |
|  | No | 866 (45.4) | 781 (41.0) | 751 (39.3) |  |
| Reasonable sleep | |  |  |  | <0.001 |
|  | Yes | 823 (43.2) | 745 (39.1) | 575 (30.1) |  |
|  | No | 1084 (56.8) | 1161 (60.9) | 1336 (69.9) |  |
| Never smoking | |  |  |  | <0.001 |
|  | Yes | 1138 (59.7) | 922 (48.4) | 883 (46.2) |  |
|  | No | 769 (40.3) | 984 (51.6) | 1028 (53.8) |  |
| No heavy alcohol consumption | |  |  |  | <0.001 |
|  | Yes | 1523 (79.9) | 1372 (72.0) | 1358 (71.1) |  |
|  | No | 384 (20.1) | 534 (28.0) | 553 (28.9) |  |
| No. of diseases | |  |  |  | <0.001 |
|  | 0 | 908 (47.6) | 828 (43.4) | 759 (39.7) |  |
|  | 1 | 505 (26.5) | 544 (28.5) | 557 (29.2) |  |
|  | 2 or more | 494 (25.9) | 534 (28.0) | 595 (31.1) |  |
| Depression | |  |  |  | <0.001 |
|  | No | 1608 (84.3) | 1490 (78.2) | 1203 (63.0) |  |
|  | Yes | 299 (15.7) | 416 (21.8) | 708 (37.1) |  |
| Childhood disadvantages | |  |  |  | <0.001 |
|  | Mild | 1369 (71.8) | 420 (22.0) | 117 (6.1) |  |
|  | Moderate | 530 (27.8) | 973 (51.1) | 403 (21.1) |  |
|  | Severe | 8 (0.4) | 513 (26.9) | 1391 (72.8) |  |
| Adulthood disadvantages | |  |  |  | <0.001 |
|  | Mild | 971 (50.9) | 557 (29.2) | 303 (15.9) |  |
|  | Moderate | 684 (35.9) | 673 (35.3) | 400 (20.9) |  |
|  | Severe | 252 (13.2) | 676 (35.5) | 1208 (63.2) |  |
| ***Childhood SES*** | |  |  |  |  |
| Parental education | |  |  |  | <0.001 |
|  | Both literate | 400 (21.0) | 284 (14.9) | 216 (11.3) |  |
|  | 1 illiterate | 730 (38.3) | 716 (37.6) | 723 (37.8) |  |
|  | Both illiterate | 777 (40.7) | 906 (47.5) | 972 (50.9) |  |
| Parental occupation | |  |  |  | <0.001 |
|  | Non-farming | 565 (29.6) | 450 (23.6) | 374 (19.6) |  |
|  | Farming | 1342 (70.4) | 1456 (76.4) | 1537 (80.4) |  |
| House type at birth | |  |  |  | <0.001 |
|  | Concrete | 387 (20.3) | 211 (11.1) | 192 (10.1) |  |
|  | Adobe | 1174 (61.6) | 1183 (62.1) | 1140 (59.7) |  |
|  | Wood or others | 346 (18.1) | 512 (26.9) | 579 (30.3) |  |
| First hukou type | |  |  |  | <0.001 |
|  | Agricultural | 1628 (85.4) | 1710 (89.7) | 1749 (91.5) |  |
|  | Non-agricultural | 279 (14.6) | 196 (10.3) | 162 (8.5) |  |
| Family’s financial situation | |  |  |  | <0.001 |
|  | A lot/somewhat better | 347 (18.2) | 189 (9.9) | 123 (6.4) |  |
|  | Same | 1239 (65.0) | 1082 (56.8) | 802 (42.0) |  |
|  | Somewhat worse/a lot worse | 321 (16.8) | 635 (33.3) | 986 (51.6) |  |
| Food deficiency | |  |  |  | <0.001 |
|  | No | 997 (52.3) | 617 (32.4) | 396 (20.7) |  |
|  | Yes | 910 (47.7) | 1289 (67.6) | 1515 (79.3) |  |
| Family starved to death | |  |  |  | <0.001 |
|  | No | 1820 (95.4) | 1754 (92.0) | 1647 (86.2) |  |
|  | Yes | 87 (4.6) | 152 (8.0) | 264 (13.8) |  |
| Parents political status | |  |  |  | <0.001 |
|  | Communist Party member | 380 (19.9) | 334 (17.5) | 285 (14.9) |  |
|  | Not Communist Party member | 1527 (80.1) | 1572 (82.5) | 1626 (85.1) |  |
| ***Childhood health*** | |  |  |  |  |
| Self-reported health status | |  |  |  | <0.001 |
|  | Healthier | 1012 (53.1) | 702 (36.8) | 519 (27.2) |  |
|  | Same | 837 (43.9) | 1027 (53.9) | 1019 (53.3) |  |
|  | Less healthy | 58 (3.0) | 177 (9.3) | 373 (19.5) |  |
| Ever confined to bed | |  |  |  | <0.001 |
|  | No | 1886 (98.9) | 1835 (96.3) | 1712 (89.6) |  |
|  | Yes | 21 (1.1) | 71 (3.7) | 199 (10.4) |  |
| Ever hospitalized for 1 or more month | |  |  |  | <0.001 |
|  | No | 1888 (99.0) | 1871 (98.2) | 1822 (95.3) |  |
|  | Yes | 19 (1.0) | 35 (1.8) | 89 (4.7) |  |
| Ever hospitalized more than 3 times | |  |  |  | <0.001 |
|  | No | 1903 (99.8) | 1897 (99.5) | 1878 (98.3) |  |
|  | Yes | 4 (0.2) | 9 (0.5) | 33 (1.7) |  |
| Received vaccinations | |  |  |  | 0.021 |
|  | Yes | 1758 (92.2) | 1709 (89.7) | 1726 (90.3) |  |
|  | No | 149 (7.8) | 197 (10.3) | 185 (9.7) |  |
| Had a usual source of care | |  |  |  | <0.001 |
|  | Yes | 1847 (96.9) | 1771 (92.9) | 1706 (89.3) |  |
|  | No | 60 (3.2) | 135 (7.1) | 205 (10.7) |  |
| ***Childhood wars*** | |  |  |  |  |
| Born in the Civil War era | |  |  |  | 0.290 |
|  | No | 1737 (91.1) | 1708 (89.6) | 1721 (90.1) |  |
|  | Yes | 170 (8.9) | 198 (10.4) | 190 (9.9) |  |
| Born in the Second Sino-Japanese era | |  |  |  |  |
|  | No | 1758 (92.2) | 1731 (90.8) | 1713 (89.6) |  |
|  | Yes | 149 (7.8) | 175 (9.2) | 198 (10.4) |  |
| ***Childhood traumas*** | |  |  |  | 0.001 |
| Parents died | | 546 (28.6) | 580 (30.4) | 651 (34.1) |  |
|  | No | 1361 (71.4) | 1326 (69.6) | 1260 (65.9) |  |
|  | Yes |  |  |  |  |
| Parents divorced | |  |  |  | 0.703 |
|  | No | 1 (0.1) | 2 (0.1) | 1 (0.1) |  |
|  | Yes | 1906 (100.0) | 1904 (99.9) | 1910 (100.0) |  |
| Sibling died | |  |  |  | <0.001 |
|  | No | 1627 (85.3) | 1498 (78.6) | 1313 (68.7) |  |
|  | Yes | 280 (14.7) | 408 (21.4) | 598 (31.3) |  |
| Parents had alcoholism problem | |  |  |  | <0.001 |
|  | No | 1818 (95.3) | 1777 (93.2) | 1718 (89.9) |  |
|  | Yes | 89 (4.7) | 129 (6.8) | 193 (10.1) |  |
| Parents had smoking problem | |  |  |  | <0.001 |
|  | No | 883 (46.3) | 822 (43.1) | 730 (38.2) |  |
|  | Yes | 1024 (53.7) | 1084 (56.9) | 1181 (61.8) |  |
| Parents had gambling problem | |  |  |  | <0.001 |
|  | No | 1894 (99.3) | 1874 (98.3) | 1856 (97.1) |  |
|  | Yes | 13 (0.7) | 32 (1.7) | 55 (2.9) |  |
| Abused by neighbor kids | |  |  |  | <0.001 |
|  | Not very often/never | 1841 (96.5) | 1719 (90.2) | 1450 (75.9) |  |
|  | Often/sometimes | 66 (3.5) | 187 (9.8) | 461 (24.1) |  |
| Abused by school kids | |  |  |  | <0.001 |
|  | Not very often/never | 1868 (98.0) | 1801 (94.5) | 1547 (81.0) |  |
|  | Often/sometimes | 39 (2.1) | 105 (5.5) | 364 (19.1) |  |
| Hit by mother | |  |  |  | <0.001 |
|  | Not very often/never | 1705 (89.4) | 1438 (75.5) | 1136 (59.5) |  |
|  | Often/sometimes | 202 (10.6) | 468 (24.6) | 775 (40.6) |  |
| Hit by father | |  |  |  | <0.001 |
|  | Not very often/never | 1805 (94.7) | 1569 (82.3) | 1282 (67.1) |  |
|  | Often/sometimes | 102 (5.4) | 337 (17.7) | 629 (32.9) |  |
| Hit by siblings | |  |  |  | <0.001 |
|  | Not very often/never | 1860 (97.5) | 1807 (94.8) | 1685 (88.2) |  |
|  | Often/sometimes | 47 (2.5) | 99 (5.2) | 226 (11.8) |  |
| Parents ever quarreled | |  |  |  | <0.001 |
|  | Not very often/never | 1724 (90.4) | 1477 (77.5) | 1117 (58.5) |  |
|  | Often/sometimes | 183 (9.6) | 429 (22.5) | 794 (41.6) |  |
| Parents hit each other | |  |  |  | <0.001 |
|  | Not very often/never | 1888 (99.0) | 1787 (93.8) | 1552 (81.2) |  |
|  | Often/sometimes | 19 (1.0) | 119 (6.2) | 359 (18.8) |  |
| Parental sadness or depression | |  |  |  | <0.001 |
|  | No | 1832 (96.1) | 1631 (85.6) | 1241 (64.9) |  |
|  | Yes | 75 (3.9) | 275 (14.4) | 670 (35.1) |  |
| Parents’ sick on bed | |  |  |  | <0.001 |
|  | No | 1760 (92.3) | 1603 (84.1) | 1388 (72.6) |  |
|  | Yes | 147 (7.7) | 303 (15.9) | 523 (27.4) |  |
| Parents’ deformity | |  |  |  | <0.001 |
|  | No | 1881 (98.6) | 1821 (95.5) | 1753 (91.7) |  |
|  | Yes | 26 (1.4) | 85 (4.5) | 158 (8.3) |  |
| Parents’ abnormality of mind | |  |  |  | <0.001 |
|  | No | 1891 (99.2) | 1867 (98.0) | 1793 (93.8) |  |
|  | Yes | 16 (0.8) | 39 (2.1) | 118 (6.2) |  |
| ***Childhood relationships*** | |  |  |  |  |
| Self-rated neighborhood safety | |  |  |  | <0.001 |
|  | Very/somewhat | 1857 (97.4) | 1787 (93.8) | 1687 (88.3) |  |
|  | Not very/not at all | 50 (2.6) | 119 (6.2) | 224 (11.7) |  |
| Self-rated neighborhood willingness to help | |  |  |  | <0.001 |
|  | Very/somewhat | 1865 (97.8) | 1796 (94.2) | 1681 (88.0) |  |
|  | Not very/not at all | 42 (2.2) | 110 (5.8) | 230 (12.0) |  |
| Self-rated neighborhood close-knit relationship | |  |  |  | <0.001 |
|  | Very/somewhat | 1896 (99.4) | 1877 (98.5) | 1814 (94.9) |  |
|  | Not very/not at all | 11 (0.6) | 29 (1.5) | 97 (5.1) |  |
| Self-rated neighborhood cleanness | |  |  |  | <0.001 |
|  | Very/somewhat | 1479 (77.6) | 1210 (63.5) | 1003 (52.5) |  |
|  | Not very/not at all | 428 (22.4) | 696 (36.5) | 908 (47.5) |  |
| Felt lonely | |  |  |  | <0.001 |
|  | Never/not very often | 1852 (97.1) | 1759 (92.3) | 1567 (82.0) |  |
|  | Sometimes/often | 55 (2.9) | 147 (7.7) | 344 (18.0) |  |
| Had a group of friends | |  |  |  | <0.001 |
|  | Sometimes/often | 1756 (92.1) | 1636 (85.8) | 1524 (79.8) |  |
|  | Never/not very often | 151 (7.9) | 270 (14.2) | 387 (20.3) |  |
| Had a good friend | |  |  |  | <0.001 |
|  | Yes | 1313 (68.9) | 1136 (59.6) | 1039 (54.4) |  |
|  | No | 594 (31.2) | 770 (40.4) | 872 (45.6) |  |
| Self-rated relationship with mother | |  |  |  | <0.001 |
|  | Excellent | 1117 (58.6) | 558 (29.3) | 335 (17.5) |  |
|  | Very good | 685 (35.9) | 727 (38.1) | 470 (24.6) |  |
|  | Good | 92 (4.8) | 412 (21.6) | 424 (22.2) |  |
|  | Fair | 13 (0.7) | 209 (11.0) | 654 (34.2) |  |
|  | Poor | 0 (0.0) | 0 (0.0) | 28 (1.5) |  |
| Mother’s love and affection | |  |  |  | <0.001 |
|  | Often/sometimes | 1719 (90.1) | 1547 (81.2) | 1365 (71.4) |  |
|  | Rarely/never | 188 (9.9) | 359 (18.8) | 546 (28.6) |  |
| Mother’s effort put into watching over you | |  |  |  | <0.001 |
|  | A lot/some | 1716 (90.0) | 1519 (79.7) | 1316 (68.9) |  |
|  | A little/not at all | 191 (10.0) | 387 (20.3) | 595 (31.1) |  |
| Mother’s preference for siblings | |  |  |  | <0.001 |
|  | A little/not at all | 1803 (94.6) | 1711 (89.8) | 1483 (77.6) |  |
|  | Very much/somewhat | 104 (5.5) | 195 (10.2) | 428 (22.4) |  |
| Mother’s preference for boy | |  |  |  | <0.001 |
|  | A little/not at all | 1862 (97.6) | 1794 (94.1) | 1627 (85.1) |  |
|  | Very much/somewhat | 45 (2.4) | 112 (5.9) | 284 (14.9) |  |
| Self-rated relationship with father | |  |  |  | <0.001 |
|  | Excellent | 1061 (55.6) | 491 (25.8) | 259 (13.6) |  |
|  | Very good | 723 (37.9) | 739 (38.8) | 466 (24.4) |  |
|  | Good | 99 (5.2) | 436 (22.9) | 411 (21.5) |  |
|  | Fair | 24 (1.3) | 237 (12.4) | 735 (38.5) |  |
|  | Poor | 0 (0.0) | 3 (0.2) | 40 (2.1) |  |
| Father’s preference for siblings | |  |  |  | <0.001 |
|  | A little/not at all | 1851 (97.1) | 1766 (92.7) | 1547 (81.0) |  |
|  | Very much/somewhat | 56 (2.9) | 140 (7.4) | 364 (19.1) |  |
| Father’s preference for boy | |  |  |  | <0.001 |
|  | A little/not at all | 1858 (97.4) | 1783 (93.6) | 1600 (83.7) |  |
|  | Very much/somewhat | 49 (2.6) | 123 (6.5) | 311 (16.3) |  |
| ***Adulthood SES*** | |  |  |  |  |
| Individual’s education attainment | |  |  |  | <0.001 |
|  | College or more | 166 (8.7) | 92 (4.8) | 69 (3.6) |  |
|  | Senior | 290 (15.2) | 200 (10.5) | 136 (7.1) |  |
|  | Middle | 676 (35.5) | 606 (31.8) | 492 (25.8) |  |
|  | Elementary | 728 (38.2) | 917 (48.1) | 1098 (57.5) |  |
|  | Illiteracy | 47 (2.5) | 91 (4.8) | 116 (6.1) |  |
| Individual’s occupation | |  |  |  | <0.001 |
|  | Others | 115 (6.0) | 91 (4.8) | 96 (5.0) |  |
|  | Non- agricultural employed | 555 (29.1) | 448 (23.5) | 352 (18.4) |  |
|  | Agricultural | 1237 (64.9) | 1367 (71.7) | 1463 (76.6) |  |
| Household income | |  |  |  | <0.001 |
|  | Third tertile | 781 (41.0) | 641 (33.6) | 492 (25.8) |  |
|  | Second tertile | 649 (34.0) | 606 (31.8) | 649 (34.0) |  |
|  | First tertile | 477 (25.0) | 659 (34.6) | 770 (40.3) |  |
| Residence | |  |  |  | <0.001 |
|  | Urban | 968 (50.8) | 801 (42.0) | 774 (40.5) |  |
|  | Rural | 939 (49.2) | 1105 (58.0) | 1137 (59.5) |  |
| ***Adulthood adversities*** | |  |  |  |  |
| Child died | |  |  |  | <0.001 |
|  | No | 1813 (95.1) | 1771 (92.9) | 1715 (89.7) |  |
|  | Yes | 94 (4.9) | 135 (7.1) | 196 (10.3) |  |
| Physically injury | |  |  |  | <0.001 |
|  | No | 1881 (98.6) | 1803 (94.6) | 1610 (84.3) |  |
|  | Yes | 26 (1.4) | 103 (5.4) | 301 (15.8) |  |
| Experienced lifetime discrimination | |  |  |  | <0.001 |
|  | No | 1873 (98.2) | 1787 (93.8) | 1584 (82.9) |  |
|  | Yes | 34 (1.8) | 119 (6.2) | 327 (17.1) |  |
| Ever confined to bed | |  |  |  | <0.001 |
|  | No | 1877 (98.4) | 1709 (89.7) | 1310 (68.6) |  |
|  | Yes | 30 (1.6) | 197 (10.3) | 601 (31.5) |  |
| Ever hospitalized for 1 or more month | |  |  |  | <0.001 |
|  | No | 1890 (99.1) | 1772 (93.0) | 1444 (75.6) |  |
|  | Yes | 17 (0.9) | 134 (7.0) | 467 (24.4) |  |
| Ever hospitalized 3 or more times | |  |  |  | <0.001 |
|  | No | 1897 (99.5) | 1866 (97.9) | 1751 (91.6) |  |
|  | Yes | 10 (0.5) | 40 (2.1) | 160 (8.4) |  |
| Ever left job for health condition | |  |  |  | <0.001 |
|  | No | 1868 (98.0) | 1668 (87.5) | 1221 (63.9) |  |
|  | Yes | 39 (2.1) | 238 (12.5) | 690 (36.1) |  |
| Had a usual source of care | |  |  |  | <0.001 |
|  | No | 1847 (96.9) | 1771 (92.9) | 1706 (89.3) |  |
|  | Yes | 60 (3.2) | 135 (7.1) | 205 (10.7) |  |

Notes: SES, socioeconomic status.

### Table S3. The mediation proportion between life-course disadvantages and depression attributed to specific lifestyles

|  | **Mediation proportion (%) (95% CI) of specific lifestyles** | | | |
| --- | --- | --- | --- | --- |
|  | **Exercise** | **Sleep** | **Smoking** | **Alcohol consumption** |
| ***Life-course disadvantages*** | |  |  |  |
| Mild | - | - | - | - |
| Moderate | **-1.7 (-4.7 to -0.1)** | **4.4 (0.6 to 9.8)** | **2.2 (0.4 to 5.3)** | 0.4 (-0.6 to 2.2) |
| Severe | **-0.8 (-1.9 to -0.1)** | **5.1 (3.3 to 7.3)** | **1.1 (0.3 to 2.2)** | 0.1 (-0.2 to 0.8) |
| ***Childhood disadvantages*** | |  |  |  |
| Mild | - | - | - | - |
| Moderate | **-1.7 (-6.0 to -0.1)** | 3.8 (-0.7 to 9.5) | 1.2 (-0.8 to 4.3) | 0 (-1.2 to 1.0) |
| Severe | **-1.2 (-2.7 to -0.2)** | **5.0 (3.0 to 7.8)** | 0.4 (-0.5 to 1.4) | 0 (-0.6 to 0.3) |
| ***Adulthood disadvantages*** | |  |  |  |
| Mild | - | - | - | - |
| Moderate | **-1.5 (-5.3 to -0.1)** | 0.7 (-4.1 to 5.9) | **2.9 (0.8 to 7.0)** | 0.1 (-0.6 to 1.5) |
| Severe | -0.2 (-1.2 to 0.3) | **2.5 (0.8 to 4.6)** | **1.9 (0.7 to 3.4)** | 0.2 (-0.2 to 0.8) |

Notes: CI, confidence interval. Models for life-course disadvantages were adjusted for age, sex, No. of diseases, and other lifestyles. Models for childhood disadvantages were adjusted for adulthood disadvantages, age, sex, No. of diseases, and other lifestyles. Models for adulthood disadvantages were adjusted for childhood disadvantages, age, sex, No. of diseases, and other lifestyles. Bolded indicates statistical significance.

### Table S4. Association between No. of healthy lifestyles and depression

| **No. of healthy lifestyles** | **Total** | **Male** | **Female** |
| --- | --- | --- | --- |
|  | **OR (95% CI)** | | |
| 0/1 | Reference | Reference | Reference |
| 2 | **0.82 (0.69-0.97)** | 0.84 (0.69-1.03) | 0.71 (0.46-1.11) |
| 3 | **0.60 (0.49-0.73)** | **0.60 (0.46-0.79)** | **0.54 (0.35-0.83)** |
| 4 | **0.34 (0.26-0.46)** | **0.26 (0.12-0.57)** | **0.31 (0.19-0.51)** |

Notes: OR, odds ratio. CI, confidence interval. All models were adjusted for age, sex, No. of diseases, and life-course disadvantages. Bolded indicates statistical significance.

### Table S5. Association between specific healthy lifestyles and depression

| **Specific healthy lifestyles** | **Total** | **Male** | **Female** |
| --- | --- | --- | --- |
|  | **OR (95% CI)** | | |
| Regular exercise | **0.87 (0.76-0.99)** | 0.87 (0.72-1.04) | 0.86 (0.72-1.03) |
| Reasonable sleep | **0.59 (0.52-0.68)** | **0.59 (0.49-0.72)** | **0.58 (0.47-0.70)** |
| Never smoking | **0.68 (0.56-0.83)** | 0.85 (0.67-1.09) | **0.48 (0.36-0.65)** |
| No heavy alcohol consumption | 0.93 (0.79-1.09) | 0.91 (0.75-1.09) | 0.91 (0.62-1.34) |

Notes: OR, odds ratio. CI, confidence interval. All models were adjusted for age, sex, No. of diseases, life-course disadvantages, and other lifestyles. Bolded indicates statistical significance.

### Table S6. Sex-stratified association of healthy lifestyles with depression by childhood and adulthood disadvantages

| **No. of healthy lifestyles** | **Mild** | **Moderate** | **Severe** | **P for interaction** |
| --- | --- | --- | --- | --- |
|  | **OR (95% CI)** | | |  |
|  | ***Life-course disadvantages*** | | |  |
| ***Male*** |  | | |  |
| 0/1 | Reference | Reference | Reference | 0.178 |
| 2 | 1.38 (0.90-2.12) | 0.86 (0.60-1.23) | **0.67 (0.50-0.89)** |  |
| 3 | 0.72 (0.40-1.28) | **0.61 (0.37-0.99)** | **0.56 (0.38-0.82)** |  |
| 4 | 0.68 (0.20-2.32) | **0.12 (0.02-0.91)** | **0.20 (0.06-0.69)** |  |
| ***Female*** |  | | |  |
| 0/1 | Reference | Reference | Reference | 0.814 |
| 2 | 0.55 (0.21-1.42) | 0.82 (0.39-1.71) | 0.71 (0.36-1.41) |  |
| 3 | **0.37 (0.15-0.95)** | 0.68 (0.33-1.41) | 0.54 (0.27-1.07) |  |
| 4 | **0.26 (0.09-0.71)** | **0.31 (0.13-0.70)** | **0.34 (0.16-0.73)** |  |
|  | ***Childhood disadvantages*** | | |  |
| ***Male*** |  | | |  |
| 0/1 | Reference | Reference | Reference | 0.291 |
| 2 | 1.22 (0.81-1.85) | 0.78 (0.55-1.11) | **0.73 (0.54-0.98)** |  |
| 3 | 0.56 (0.30-1.01) | 0.72 (0.46-1.12) | **0.53 (0.35-0.80)** |  |
| 4 | 0.58 (0.17-1.97) | 0.32 (0.07-1.41) | **0.13 (0.03-0.55)** |  |
| ***Female*** |  | | |  |
| 0/1 | Reference | Reference | Reference | 0.300 |
| 2 | 0.55 (0.22-1.39) | 0.62 (0.30-1.29) | 1.00 (0.49-2.06) |  |
| 3 | **0.29 (0.11-0.74)** | 0.61 (0.30-1.23) | 0.78 (0.38-1.60) |  |
| 4 | **0.21 (0.08-0.57)** | **0.30 (0.14-0.68)** | 0.45 (0.20-1.01) |  |
|  | ***Adulthood disadvantages*** | | |  |
| ***Male*** |  | | |  |
| 0/1 | Reference | Reference | Reference | 0.420 |
| 2 | 0.93 (0.61-1.41) | 0.90 (0.62-1.32) | 0.77 (0.58-1.02) |  |
| 3 | 0.59 (0.34-1.05) | **0.48 (0.28-0.81)** | **0.71 (0.49-1.03)** |  |
| 4 | 0.62 (0.21-1.83) | 0.28 (0.06-1.23) | **0.08 (0.01-0.58)** |  |
| ***Female*** |  | | |  |
| 0/1 | Reference | Reference | Reference | 0.194 |
| 2 | 0.53 (0.19-1.50) | 1.37 (0.56-3.36) | 0.59 (0.32-1.11) |  |
| 3 | 0.48 (0.17-1.35) | 0.85 (0.35-2.07) | **0.47 (0.25-0.88)** |  |
| 4 | **0.16 (0.05-0.51)** | 0.73 (0.28-1.88) | **0.27 (0.13-0.55)** |  |

Notes: OR, odds ratio. CI, confidence interval. Models for life-course disadvantages were adjusted for age and No. of diseases. Models for childhood disadvantages were adjusted for adulthood disadvantages, age, and No. of diseases. Models for adulthood disadvantages were adjusted for childhood disadvantages, age, and No. of diseases. Bolded indicates statistical significance.

### Table S7. Sex-stratified association of specific healthy lifestyles with depression by childhood and adulthood disadvantages

| **Specific healthy lifestyles** | **Mild** | **Moderate** | **Severe** | **P for interaction** |
| --- | --- | --- | --- | --- |
|  | **OR (95% CI)** | | |  |
|  | ***Life-course disadvantages*** | | |  |
| ***Male*** |  | | |  |
| Regular exercise | 1.47 (0.98-2.19) | **0.71 (0.51-0.99)** | **0.75 (0.58-0.99)** | 0.021 |
| Reasonable sleep | **0.57 (0.38-0.85)** | 0.74 (0.53-1.05) | **0.51 (0.39-0.69)** | 0.297 |
| Never smoking | 0.65 (0.39-1.09) | 0.96 (0.62-1.50) | 0.91 (0.64-1.29) | 0.640 |
| No heavy alcohol consumption | 1.17 (0.78-1.74) | 0.73 (0.53-1.02) | 0.90 (0.69-1.18) | 0.268 |
| ***Female*** |  | | |  |
| Regular exercise | 0.90 (0.63-1.29) | 0.91 (0.66-1.24) | 0.80 (0.60-1.07) | 0.813 |
| Reasonable sleep | **0.51 (0.35-0.74)** | **0.53 (0.38-0.74)** | **0.70 (0.51-0.97)** | 0.362 |
| Never smoking | **0.54 (0.30-0.97)** | **0.49 (0.28-0.86)** | **0.43 (0.27-0.70)** | 0.833 |
| No heavy alcohol consumption | 0.78 (0.35-1.77) | 0.83 (0.44-1.59) | 1.04 (0.57-1.86) | 0.574 |
|  | ***Childhood disadvantages*** | | |  |
| ***Male*** |  | | |  |
| Regular exercise | 1.33 (0.90-1.95) | 0.78 (0.57-1.08) | **0.72 (0.54-0.96)** | 0.100 |
| Reasonable sleep | **0.58 (0.39-0.87)** | **0.66 (0.47-0.92)** | **0.56 (0.41-0.75)** | 0.723 |
| Never smoking | 0.80 (0.48-1.34) | 0.96 (0.63-1.47) | 0.82 (0.57-1.18) | 0.761 |
| No heavy alcohol consumption | 0.95 (0.64-1.39) | 0.95 (0.69-1.31) | 0.85 (0.64-1.12) | 0.848 |
| ***Female*** |  | | |  |
| Regular exercise | 0.75 (0.52-1.07) | 0.95 (0.69-1.31) | 0.83 (0.61-1.13) | 0.874 |
| Reasonable sleep | **0.44 (0.30-0.65)** | **0.64 (0.45-0.90)** | **0.62 (0.44-0.86)** | 0.342 |
| Never smoking | **0.54 (0.30-0.97)** | **0.54 (0.32-0.91)** | **0.53 (0.32-0.87)** | 0.987 |
| No heavy alcohol consumption | 0.75 (0.35-1.61) | 0.74 (0.37-1.51) | 1.35 (0.74-2.46) | 0.247 |
|  | ***Adulthood disadvantages*** | | |  |
| ***Male*** |  | | |  |
| Regular exercise | 0.91 (0.62-1.34) | 0.86 (0.60-1.24) | 0.84 (0.64-1.09) | 0.988 |
| Reasonable sleep | **0.57 (0.38-0.86)** | **0.60 (0.42-0.86)** | **0.62 (0.47-0.82)** | 0.910 |
| Never smoking | 0.72 (0.45-1.17) | 0.84 (0.52-1.37) | 0.97 (0.69-1.38) | 0.605 |
| No heavy alcohol consumption | 1.15 (0.78-1.70) | 0.79 (0.56-1.13) | 0.85 (0.65-1.10) | 0.389 |
| ***Female*** |  | | |  |
| Regular exercise | 0.90 (0.61-1.33) | 0.74 (0.54-1.02) | 0.92 (0.70-1.23) | 0.379 |
| Reasonable sleep | **0.38 (0.24-0.59)** | 0.75 (0.54-1.06) | **0.56 (0.41-0.76)** | 0.072 |
| Never smoking | **0.41 (0.20-0.84)** | 0.96 (0.52-1.76) | **0.40 (0.26-0.62)** | 0.070 |
| No heavy alcohol consumption | 2.48 (0.72-8.55) | 0.57 (0.29-1.13) | 1.06 (0.61-1.83) | 0.215 |

Notes: OR, odds ratio. CI, confidence interval. Models for life-course disadvantages were adjusted for age, No. of diseases, and other lifestyles. Models for childhood disadvantages were adjusted for adulthood disadvantages, age, No. of diseases, and other lifestyles. Models for adulthood disadvantages were adjusted for childhood disadvantages, age, No. of diseases, and other lifestyles. Bolded indicates statistical significance.

### Table S8. Sex-stratified joint association of life-course disadvantages and unhealthy lifestyles with depression

| **No. of healthy lifestyles** | **Life-course disadvantages** | **Childhood disadvantages** | **Adulthood disadvantages** |
| --- | --- | --- | --- |
|  | **OR (95% CI)** | | |
|  | ***Male*** | | |
| ***Mild*** | | | |
| 4 | Reference | Reference | Reference |
| 3 | 0.99 (0.27-3.61) | 0.86 (0.23-3.23) | 0.98 (0.31-3.05) |
| 2 | 2.13 (0.63-7.16) | 1.75 (0.51-5.95) | 1.55 (0.53-4.53) |
| 0/1 | 1.51 (0.44-5.13) | 1.48 (0.44-5.03) | 1.70 (0.58-4.95) |
| ***Moderate*** |  |  |  |
| 4 | 0.40 (0.04-4.04) | 0.86 (0.13-5.60) | 0.61 (0.10-3.59) |
| 3 | 1.80 (0.52-6.25) | 2.02 (0.59-6.97) | 1.07 (0.35-3.29) |
| 2 | 2.66 (0.80-8.83) | 2.24 (0.67-7.52) | 1.97 (0.68-5.72) |
| 0/1 | 2.95 (0.89-9.77) | 2.99 (0.90-10.00) | 2.20 (0.76-6.39) |
| ***Severe*** |  |  |  |
| 4 | 1.54 (0.29-8.27) | 0.67 (0.10-4.29) | 0.30 (0.03-2.87) |
| 3 | **4.47 (1.33-15.06)** | 2.93 (0.86-10.04) | 2.79 (0.95-8.24) |
| 2 | **5.20 (1.58-17.11)** | **4.13 (1.24-13.74)** | **3.05 (1.06-8.75)** |
| 0/1 | **7.60 (2.32-24.92)** | **5.22 (1.57-17.35)** | **3.93 (1.38-11.23)** |
|  | ***Female*** | | |
| ***Mild*** | | | |
| 4 | Reference | Reference | Reference |
| 3 | 1.65 (0.99-2.74) | 1.33 (0.83-2.15) | **3.06 (1.58-5.96)** |
| 2 | **2.81 (1.66-4.75)** | **2.36 (1.44-3.86)** | **3.33 (1.66-6.66)** |
| 0/1 | **4.58 (1.76-11.94)** | **4.31 (1.71-10.88)** | **6.24 (1.96-19.88)** |
| ***Moderate*** |  |  |  |
| 4 | 1.64 (0.89-3.02) | 1.22 (0.67-2.21) | **3.74 (1.80-7.76)** |
| 3 | **3.46 (2.08-5.74)** | **2.17 (1.35-3.47)** | **4.39 (2.27-8.48)** |
| 2 | **3.37 (1.97-5.76)** | **2.47 (1.51-4.06)** | **7.21 (3.69-14.08)** |
| 0/1 | **5.02 (2.23-11.28)** | **3.15 (1.45-6.85)** | **5.12 (1.78-14.74)** |
| ***Severe*** |  |  |  |
| 4 | **3.85 (2.06-7.17)** | 1.77 (0.96-3.27) | **5.16 (2.49-10.70)** |
| 3 | **5.49 (3.32-9.07)** | **3.66 (2.28-5.89)** | **9.10 (4.75-17.43)** |
| 2 | **7.47 (4.45-12.53)** | **4.31 (2.63-7.07)** | **11.45 (5.91-22.16)** |
| 0/1 | **9.71 (4.26-22.13)** | **4.84 (2.06-11.36)** | **18.77 (8.01-43.97)** |

Notes: OR, odds ratio. CI, confidence interval. Models for life-course disadvantages were adjusted for age and No. of diseases. Models for childhood disadvantages were adjusted for adulthood disadvantages, age, and No. of diseases. Models for adulthood disadvantages were adjusted for childhood disadvantages, age, and No. of diseases. Bolded indicates statistical significance.

### Table S9. The sex-stratified dilution effect of No. of healthy lifestyles on the association between life-course disadvantages and depression

| **Life-course disadvantages and No. of healthy lifestyles** | **Life-course disadvantages** | **Childhood disadvantages** | **Adulthood disadvantages** |
| --- | --- | --- | --- |
|  | **OR (95% CI)** | | |
|  | ***Male*** | | |
| Severe and 4 healthy lifestyles | Reference | Reference | Reference |
| Severe and 3 healthy lifestyles | 2.90 (0.84-10.07) | **4.40 (1.01-19.26)** | **9.28 (1.22-70.69)** |
| Severe and 2 healthy lifestyles | 3.37 (0.99-11.44) | **6.20 (1.45-26.46)** | **10.13 (1.35-75.99)** |
| Severe and 0/1 healthy lifestyles | **4.93 (1.46-16.65)** | **7.83 (1.84-33.38)** | **13.06 (1.75-97.78)** |
| Moderate and 4 healthy lifestyles | 0.26 (0.03-2.66) | 1.30 (0.17-9.96) | 2.01 (0.17-23.87) |
| Moderate and 3 healthy lifestyles | 1.17 (0.33-4.18) | 3.03 (0.69-13.36) | 3.55 (0.46-27.66) |
| Moderate and 2 healthy lifestyles | 1.73 (0.50-5.90) | 3.35 (0.78-14.46) | 6.55 (0.87-49.46) |
| Moderate and 0/1 healthy lifestyle | 1.92 (0.56-6.53) | **4.49 (1.05-19.23)** | 7.30 (0.97-55.19) |
| Mild and 4 healthy lifestyles | 0.65 (0.12-3.48) | 1.50 (0.23-9.65) | 3.32 (0.35-31.66) |
| Mild and 3 healthy lifestyles | 0.64 (0.17-2.41) | 1.30 (0.27-6.12) | 3.24 (0.41-25.50) |
| Mild and 2 healthy lifestyles | 1.38 (0.40-4.79) | 2.62 (0.60-11.45) | 5.14 (0.68-39.03) |
| Mild and 0/1 healthy lifestyle | 0.98 (0.28-3.43) | 2.22 (0.51-9.67) | 5.64 (0.74-42.71) |
|  | ***Female*** | | |
| Severe and 4 healthy lifestyles | Reference | Reference | Reference |
| Severe and 3 healthy lifestyles | 1.43 (0.89-2.30) | **2.07 (1.25-3.42)** | **1.77 (1.14-2.73)** |
| Severe and 2 healthy lifestyles | **1.94 (1.19-3.17)** | **2.44 (1.45-4.10)** | **2.22 (1.41-3.49)** |
| Severe and 0/1 healthy lifestyle | **2.52 (1.13-5.66)** | **2.74 (1.15-6.51)** | **3.64 (1.80-7.34)** |
| Moderate and 4 healthy lifestyles | **0.43 (0.24-0.77)** | 0.69 (0.37-1.28) | 0.73 (0.42-1.26) |
| Moderate and 3 healthy lifestyles | 0.90 (0.56-1.45) | 1.22 (0.74-2.02) | 0.85 (0.54-1.34) |
| Moderate and 2 healthy lifestyles | 0.88 (0.53-1.46) | 1.40 (0.83-2.36) | 1.40 (0.87-2.23) |
| Moderate and 0/1 healthy lifestyle | 1.31 (0.59-2.89) | 1.78 (0.81-3.93) | 0.99 (0.39-2.54) |
| Mild and 4 healthy lifestyles | **0.26 (0.14-0.49)** | 0.57 (0.31-1.04) | **0.19 (0.09-0.40)** |
| Mild and 3 healthy lifestyles | **0.43 (0.26-0.70)** | 0.75 (0.45-1.25) | **0.59 (0.37-0.95)** |
| Mild and 2 healthy lifestyles | 0.73 (0.44-1.20) | 1.33 (0.79-2.25) | 0.65 (0.39-1.07) |
| Mild and 0/1 healthy lifestyle | 1.19 (0.46-3.06) | 2.44 (0.95-6.25) | 1.21 (0.42-3.49) |

Notes: OR, odds ratio. CI, confidence interval. Models for life-course disadvantages were adjusted for age and No. of diseases. Models for childhood disadvantages were adjusted for adulthood disadvantages, age, and No. of diseases. Models for adulthood disadvantages were adjusted for childhood disadvantages, age, and No. of diseases. Bolded indicates statistical significance.

### Table S10. The dilution effect of specific healthy lifestyles on the association between life-course disadvantages and depression

| **Life-course disadvantages and No. of healthy lifestyles** | **Life-course disadvantages** | **Childhood disadvantages** | **Adulthood disadvantages** |
| --- | --- | --- | --- |
|  | **OR (95% CI)** | | |
| **Regular exercise** |  |  |  |
| Severe and regular exercise | Reference | Reference | Reference |
| Severe and no regular exercise | **1.27 (1.04-1.55)** | **1.26 (1.03-1.54)** | 1.13 (0.94-1.37) |
| Moderate and regular exercise | **0.51 (0.42-0.62)** | **0.62 (0.51-0.75)** | **0.54 (0.44-0.66)** |
| Moderate and no regular exercise | **0.56 (0.45-0.69)** | **0.72 (0.59-0.89)** | **0.68 (0.55-0.85)** |
| Mild and regular exercise | **0.34 (0.27-0.42)** | **0.45 (0.36-0.56)** | **0.38 (0.31-0.48)** |
| Mild and no regular exercise | **0.35 (0.28-0.44)** | **0.47 (0.37-0.59)** | **0.42 (0.33-0.54)** |
| **Reasonable sleep** |  |  |  |
| Severe and reasonable sleep | Reference | Reference | Reference |
| Severe and no reasonable sleep | **1.60 (1.29-1.99)** | **1.66 (1.33-2.07)** | **1.69 (1.38-2.07)** |
| Moderate and reasonable sleep | **0.47 (0.36-0.62)** | **0.62 (0.47-0.81)** | **0.62 (0.48-0.81)** |
| Moderate and no reasonable sleep | **0.77 (0.62-0.97)** | 0.98 (0.78-1.24) | 0.91 (0.73-1.14) |
| Mild and reasonable sleep | **0.27 (0.21-0.37)** | **0.38 (0.28-0.51)** | **0.32 (0.24-0.44)** |
| Mild and no reasonable sleep | **0.52 (0.41-0.66)** | **0.72 (0.56-0.92)** | **0.68 (0.54-0.86)** |
| **Never smoking** |  |  |  |
| Severe and never smoking | Reference | Reference | Reference |
| Severe and ever smoking | **1.56 (1.22-1.98)** | **1.42 (1.11-1.81)** | **1.34 (1.06-1.69)** |
| Moderate and never smoking | **0.52 (0.42-0.64)** | **0.60 (0.49-0.74)** | **0.59 (0.48-0.72)** |
| Moderate and ever smoking | **0.69 (0.53-0.89)** | 0.86 (0.66-1.11) | **0.72 (0.55-0.93)** |
| Mild and never smoking | **0.32 (0.26-0.39)** | **0.43 (0.35-0.54)** | **0.33 (0.27-0.42)** |
| Mild and ever smoking | **0.48 (0.36-0.63)** | **0.55 (0.42-0.74)** | **0.60 (0.45-0.79)** |
| **No heavy alcohol consumption** |  |  |  |
| Severe and no heavy alcohol consumption | Reference | Reference | Reference |
| Severe and heavy alcohol consumption | 1.11 (0.89-1.38) | 1.09 (0.87-1.37) | 1.05 (0.84-1.30) |
| Moderate and no heavy alcohol consumption | **0.48 (0.41-0.58)** | **0.59 (0.50-0.70)** | **0.54 (0.46-0.65)** |
| Moderate and heavy alcohol consumption | **0.53 (0.41-0.68)** | **0.69 (0.53-0.89)** | **0.67 (0.51-0.87)** |
| Mild and no heavy alcohol consumption | **0.32 (0.26-0.38)** | **0.43 (0.35-0.51)** | **0.38 (0.31-0.46)** |
| Mild and heavy alcohol consumption | **0.31 (0.23-0.44)** | **0.41 (0.29-0.56)** | **0.38 (0.27-0.52)** |

Notes: OR, odds ratio. CI, confidence interval. Models for life-course disadvantages were adjusted for age, sex, No. of diseases, and other lifestyles. Models for childhood disadvantages were adjusted for adulthood disadvantages, age, sex, No. of diseases, and other lifestyles. Models for adulthood disadvantages were adjusted for childhood disadvantages, age, sex, No. of diseases, and other lifestyles. Bolded indicates statistical significance.

### Table S11. The sex-stratified dilution effect of specific healthy lifestyles on the association between life-course disadvantages and depression

| **Life-course disadvantages and No. of healthy lifestyles** | **Life-course disadvantages** | **Childhood disadvantages** | **Adulthood disadvantages** |
| --- | --- | --- | --- |
|  | **OR (95% CI)** | | |
|  | ***Male*** | | |
| **Regular exercise** |  |  |  |
| Severe and regular exercise | Reference | Reference | Reference |
| Severe and no regular exercise | **1.32 (1.02-1.71)** | 1.28 (0.99-1.67) | 1.18 (0.91-1.53) |
| Moderate and regular exercise | **0.46 (0.35-0.60)** | **0.60 (0.46-0.79)** | **0.57 (0.44-0.76)** |
| Moderate and no regular exercise | **0.54 (0.41-0.73)** | 0.75 (0.56-1.01) | **0.68 (0.49-0.95)** |
| Mild and regular exercise | **0.36 (0.26-0.49)** | **0.44 (0.32-0.61)** | **0.47 (0.34-0.64)** |
| Mild and no regular exercise | **0.28 (0.19-0.40)** | **0.34 (0.24-0.50)** | **0.53 (0.38-0.73)** |
| **Reasonable sleep** |  |  |  |
| Severe and reasonable sleep | Reference | Reference | Reference |
| Severe and no reasonable sleep | **1.78 (1.35-2.36)** | **1.71 (1.29-2.20)** | **1.62 (1.23-2.14)** |
| Moderate and reasonable sleep | **0.49 (0.34-0.70)** | **0.62 (0.44-0.89)** | **0.56 (0.39-0.82)** |
| Moderate and no reasonable sleep | 0.75 (0.55-1.01) | 1.00 (0.74-1.36) | 0.94 (0.68-1.29) |
| Mild and reasonable sleep | **0.29 (0.19-0.45)** | **0.35 (0.23-0.54)** | **0.43 (0.28-0.65)** |
| Mild and no reasonable sleep | **0.50 (0.35-0.71)** | **0.60 (0.42-0.86)** | 0.77 (0.55-1.07) |
| **Never smoking** |  |  |  |
| Severe and never smoking | Reference | Reference | Reference |
| Severe and ever smoking | 1.13 (0.80-1.58) | 1.14 (0.81-1.60) | 1.05 (0.74-1.48) |
| Moderate and never smoking | **0.44 (0.27-0.73)** | **0.56 (0.34-0.93)** | **0.51 (0.30-0.88)** |
| Moderate and ever smoking | **0.49 (0.35-0.70)** | **0.69 (0.48-0.98)** | **0.61 (0.42-0.89)** |
| Mild and never smoking | **0.23 (0.13-0.41)** | **0.37 (0.21-0.66)** | **0.37 (0.22-0.63)** |
| Mild and ever smoking | **0.34 (0.23-0.49)** | **0.39 (0.26-0.58)** | **0.50 (0.34-0.74)** |
| **No heavy alcohol consumption** |  |  |  |
| Severe and no heavy alcohol consumption | Reference | Reference | Reference |
| Severe and heavy alcohol consumption | 1.09 (0.85-1.41) | 1.14 (0.88-1.47) | 1.15 (0.89-1.49) |
| Moderate and no heavy alcohol consumption | **0.41 (0.31-0.54)** | **0.59 (0.45-0.78)** | **0.55 (0.41-0.74)** |
| Moderate and heavy alcohol consumption | **0.52 (0.39-0.69)** | **0.68 (0.51-0.91)** | **0.69 (0.51-0.94)** |
| Mild and no heavy alcohol consumption | **0.31 (0.23-0.42)** | **0.37 (0.27-0.52)** | **0.51 (0.38-0.69)** |
| Mild and heavy alcohol consumption | **0.27 (0.18-0.40)** | **0.37 (0.26-0.54)** | **0.45 (0.32-0.64)** |
|  | ***Female*** | | |
| **Regular exercise** |  |  |  |
| Severe and regular exercise | Reference | Reference | Reference |
| Severe and no regular exercise | 1.23 (0.91-1.66) | 1.22 (0.89-1.69) | 1.08 (0.82-1.42) |
| Moderate and regular exercise | **0.59 (0.44-0.78)** | **0.63 (0.47-0.84)** | **0.49 (0.37-0.66)** |
| Moderate and no regular exercise | **0.59 (0.43-0.80)** | **0.70 (0.51-0.95)** | **0.69 (0.51-0.93)** |
| Mild and regular exercise | **0.33 (0.25-0.44)** | **0.47 (0.35-0.64)** | **0.31 (0.22-0.42)** |
| Mild and no regular exercise | **0.42 (0.31-0.57)** | **0.58 (0.42-0.79)** | **0.33 (0.23-0.46)** |
| **Reasonable sleep** |  |  |  |
| Severe and reasonable sleep | Reference | Reference | Reference |
| Severe and no reasonable sleep | 1.38 (0.98-1.94) | **1.60 (1.12-2.29)** | **1.77 (1.31-2.40)** |
| Moderate and reasonable sleep | **0.45 (0.30-0.67)** | **0.61 (0.40-0.93)** | **0.68 (0.46-0.99)** |
| Moderate and no reasonable sleep | 0.79 (0.56-1.12) | 0.96 (0.67-1.38) | 0.90 (0.66-1.24) |
| Mild and reasonable sleep | **0.25 (0.17-0.38)** | **0.39 (0.26-0.60)** | **0.23 (0.14-0.36)** |
| Mild and no reasonable sleep | **0.51 (0.36-0.73)** | 0.82 (0.56-1.18) | **0.60 (0.43-0.84)** |
| **Never smoking** |  |  |  |
| Severe and never smoking | Reference | Reference | Reference |
| Severe and ever smoking | **2.26 (1.37-3.74)** | **2.02 (1.17-3.50)** | **2.46 (1.60-3.79)** |
| Moderate and never smoking | **0.55 (0.44-0.69)** | **0.62 (0.49-0.78)** | **0.60 (0.48-0.75)** |
| Moderate and ever smoking | 1.03 (0.60-1.76) | 0.96 (0.57-1.60) | 0.61 (0.34-1.11) |
| Mild and never smoking | **0.34 (0.27-0.43)** | **0.47 (0.37-0.59)** | **0.31 (0.24-0.40)** |
| Mild and ever smoking | 0.69 (0.40-1.18) | 1.02 (0.60-1.74) | 0.69 (0.34-1.38) |
| **No heavy alcohol consumption** |  |  |  |
| Severe and no heavy alcohol consumption | Reference | Reference | Reference |
| Severe and heavy alcohol consumption | 0.95 (0.52-1.73) | 0.82 (0.44-1.53) | 0.98 (0.57-1.68) |
| Moderate and no heavy alcohol consumption | **0.54 (0.43-0.67)** | **0.59 (0.47-0.74)** | **0.54 (0.43-0.67)** |
| Moderate and heavy alcohol consumption | **0.47 (0.24-0.92)** | 0.72 (0.36-1.43) | 0.91 (0.46-1.80) |
| Mild and no heavy alcohol consumption | **0.33 (0.26-0.41)** | **0.46 (0.36-0.58)** | **0.31 (0.24-0.40)** |
| Mild and heavy alcohol consumption | 0.56 (0.27-1.16) | 0.55 (0.27-1.12) | **0.14 (0.04-0.47)** |

Notes: OR, odds ratio. CI, confidence interval. Models for life-course disadvantages were adjusted for age, No. of diseases, and other lifestyles. Models for childhood disadvantages were adjusted for adulthood disadvantages, age, No. of diseases, and other lifestyles. Models for adulthood disadvantages were adjusted for childhood disadvantages, age, No. of diseases, and other lifestyles. Bolded indicates statistical significance.

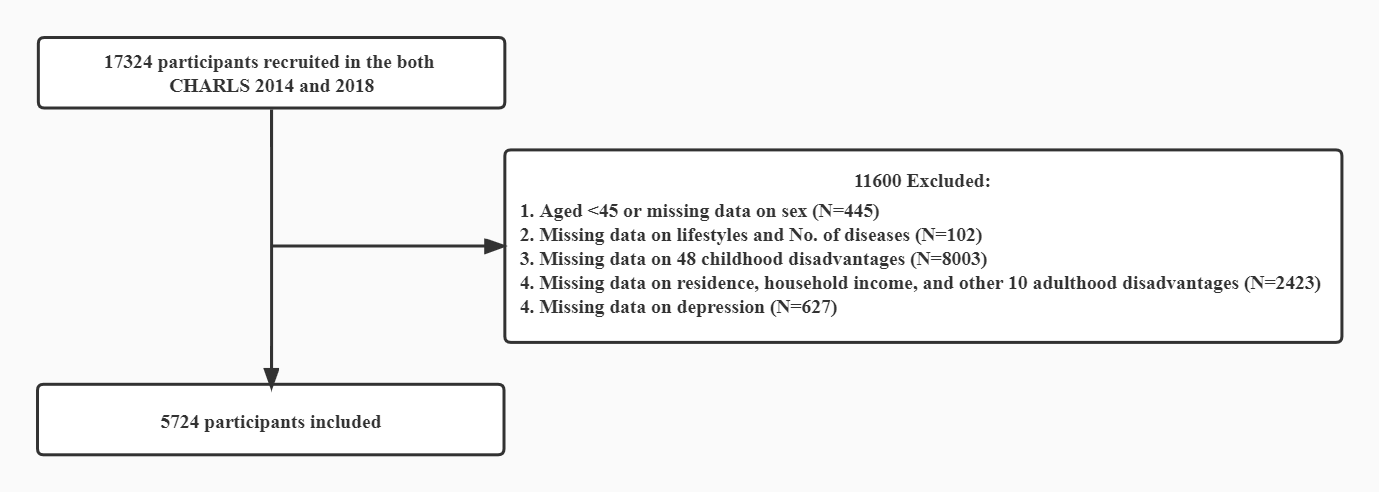

### Figure S1. Flow Chart

Notes: CHARLS, China Health and Retirement Longitudinal Study.

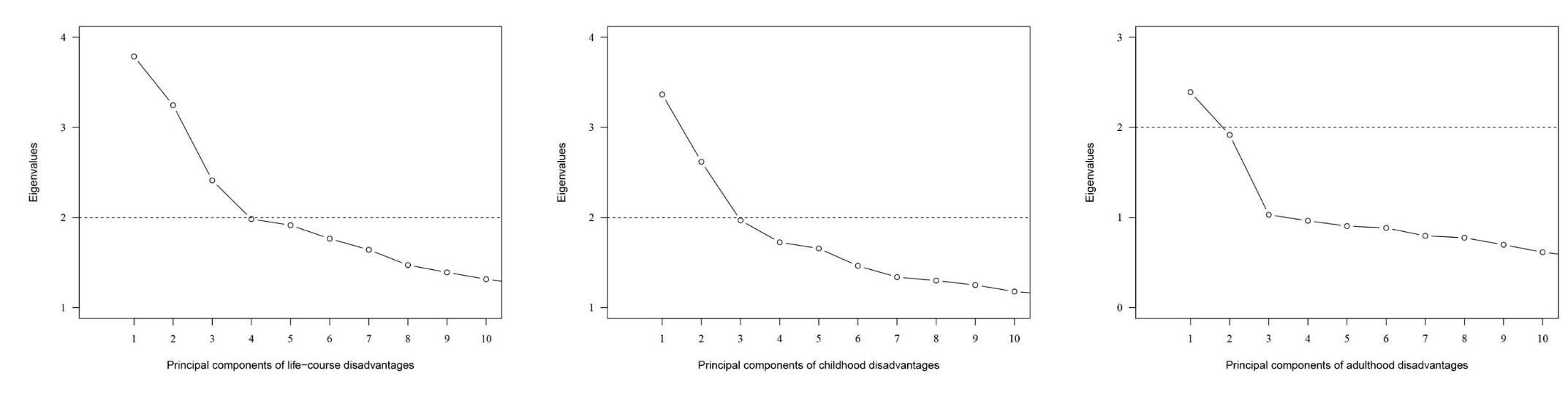

### Figure S2. Eigenvalues of principal component number

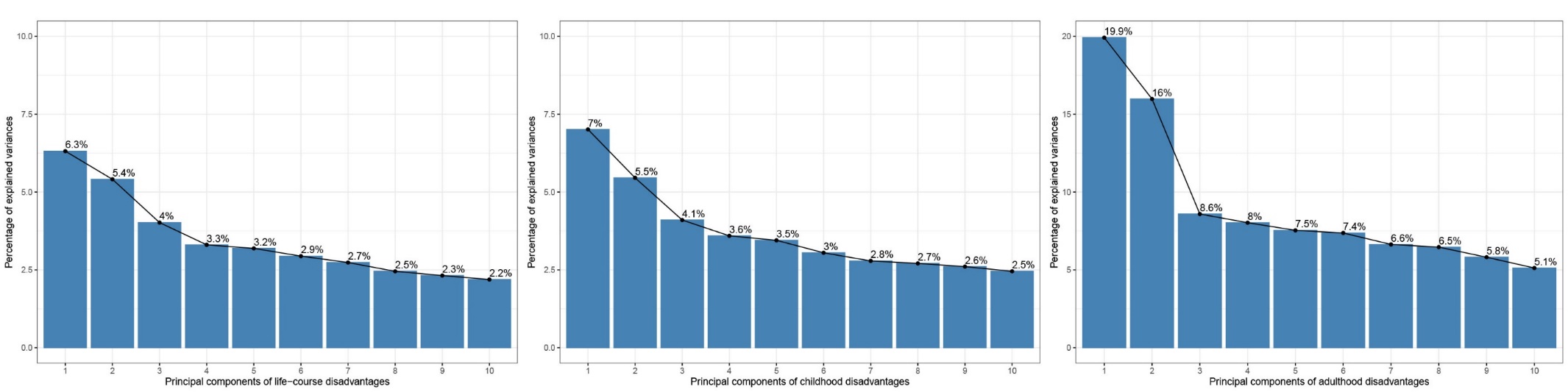

### Figure S3. Percentage of explained variances of principal component number
